# Alzheimer’s disease genetic risk and sleep phenotypes: association with more slow-waves and daytime sleepiness

**DOI:** 10.1101/2020.02.26.20027912

**Authors:** Vincenzo Muto, Ekaterina Koshmanova, Pouya Ghaemmaghami, Mathieu Jaspar, Christelle Meyer, Mahmoud Elansary, Maxime Van Egroo, Daphne Chylinski, Christian Berthomier, Marie Brandewinder, Charlotte Mouraux, Christina Schmidt, Grégory Hammad, Wouter Coppieters, Naima Ahariz, Christian Degueldre, André Luxen, Eric Salmon, Christophe Phillips, Simon N. Archer, Loic Yengo, Enda Byrne, Fabienne Collette, Michel Georges, Derk-Jan Dijk, Pierre Maquet, Peter M. Visscher, Gilles Vandewalle

**Affiliations:** GIGA-Cyclotron Research Centre-In Vivo Imaging, University of Liège, Liège, Belgium; Walloon Excellence in Life sciences and Biotechnology (WELBIO, Belgium); Psychology and Cognitive Neuroscience Research Unit, University of Liège, Liège, Belgium; GIGA-Medical Genomics, University of Liège, Liège, Belgium; Physip, Paris, France; Department of Neurology, University Hospital of Liège, Liège, Belgium; GIGA-In Silico Medicine, University of Liège, Liège, Belgium; Sleep Research Centre University of Surrey, University of Surrey, Guildford; UK Dementia Research Institute at the University of Surrey; Institute for Molecular Bioscience, University of Queensland, Brisbane, Australia

**Keywords:** Alzheimer’s disease, polygenic risk scores, slow wave energy, daytime sleepiness

## Abstract

**Study Objectives:** Sleep disturbances and genetic variants have been identified as risk factors for Alzheimer’s disease. Our goal was to assess whether genome-wide polygenic risk scores (PRS) for AD associate with sleep phenotypes in young adults, decades before typical AD symptom onset.

**Methods:** We computed whole-genome Polygenic Risk Scores (PRS) for AD and extensively phenotyped sleep under different sleep conditions, including baseline sleep, recovery sleep following sleep deprivation and extended sleep opportunity, in a carefully selected homogenous sample of healthy 363 young men (22.1 y ± 2.7) devoid of sleep and cognitive disorders.

**Results:** AD PRS was associated with more slow wave energy, i.e. the cumulated power in the 0.5-4 Hz EEG band, a marker of sleep need, during habitual sleep and following sleep loss, and potentially with large slow wave sleep rebound following sleep deprivation. Furthermore, higher AD PRS was correlated with higher habitual daytime sleepiness.

**Conclusions:** These results imply that sleep features may be associated with AD liability in young adults, when current AD biomarkers are typically negative, and the notion that quantifying sleep alterations may be useful in assessing the risk for developing AD.

**Statement of Significance:** We show that the genetic liability for developing for Alzheimer’s disease (AD), as grasped over the entire genome using polygenic risk scores (PRS), is associated with sleep intensity and daytime sleepiness in healthy individuals devoid of sleep disorders and aged < 30 y, i.e. 30 to 60 years before typical onset of AD cognitive symptoms. Sleep features may be associated with AD liability in young adults, when current AD biomarkers are typically negative. The findings reinforce the notion that quantifying sleep alterations may be useful in assessing the risk for developing AD.

## Introduction

Defective proteostasis of brain amyloid-beta (Aβ) and tau protein antedates the clinical manifestations of Alzheimer’s disease by decades ^1–3^. This so-called “preclinical” window constitutes an opportunity for internvention that would hopefully reduce the predicted increase in AD prevalence ^4^, despite the absence of disease modifying treatments in the foreseeable future. In this respect, the further identification of AD risk factors is of paramount importance.

AD patients can become restless at night and sleepy during daytime while their entire sleep-wake cycle becomes fragmented and disorganized ^5^. Critically, similarly to Rapid Eye Movement (REM) sleep behavioral disorder (RBD) in Parkinson’s disease ^6^, altered sleep has recently been related to increased risk for developing AD, over and above sleep disturbances in AD patients ^5^. Longer latency to fall asleep and reduced sleep slow waves and rapid eye movement (REM) sleep are associated with both Aβ plaques and Tau neurofibrillary tangles (NFTs) in cognitively normal participants ^7–9^. Sleep fragmentation and the reduction in REM sleep quantity in cognitively normal individuals aged >60 y predict the future risk of developing AD ^10,11^. Acute sleep deprivation ^12,13^, and experimentally induced reduction of sleep slow waves ^14^, increases cerebrospinal fluid (CSF) Aβ and Tau protein content.

In *post mortem* human brain tissues, the first signs of brain protein aggregation are identified in the locus coeruleus (LC), a brainstem nucleus essential to sleep regulation ^15^, under the form of pretangles, consisting of phosphorylated Tau protein ^16^. Critically, LC pretangles can be detected during adolescence, while by age 30, they can be detected in the majority of the population (> 90%) ^16^. With age, Tau deposits increase in the brain in a stereotypical manner and are tightly associated with cognitive decline in overt ‘clinical’ AD ^16^. Individual variations in these intrinsic properties should be reflected in brain function, including sleep, whether or not Tau aggregation has already occurred.

Sporadic AD, the most common form of AD in the general population, has an estimated heritability ranging between 58% to 79% ^17,18^. Individual Polygenic Risk Scores (PRS) for AD can be computed based on results of published Genome Wide Association Studies (GWAS). These PRS reflect part of the genetic liability for AD in any asymptomatic individual and, at the group level, can be associated with phenotypes of interest which are related to the (risk) pathways leading to AD ^19,20^. Recent studies reported significant association between AD PRS and CSF Aβ content ^21,22^, cortical thickness ^23^, memory decline ^24^, and hippocampus volume ^22,25,26^ in cognitively normal older adults (> 45 y) but, importantly, also in young adults (18 - 35 y) ^25^.

Here, we conducted a proof-of-concept study to establish that sleep can be related to AD risk in young adults, using PRS for AD. We phenotyped sleep under different conditions (baseline, sleep extension, recovery sleep after total sleep deprivation) in a homogenous sample of young healthy cognitively normal men without sleep disorders and computed individual PRS for AD. We hypothesized that high PRS would be associated with sleep metrics that had previously been associated with AD features in cognitively normal older adults. We further explored whether subjective assessments and behavioural correlates of sleep quality would be associated with PRS for AD.

## Methods

This research was approved by the Ethics Committee of the Faculty of Medicine at the University of Liège, Belgium.

### Participants

All participants signed an informed consent prior to their participation and received a financial compensation. Three hundred and sixty-four young healthy men (aged 18-31 years) were enrolled for the study after giving their written informed consent, and received a financial compensation. Exclusion criteria were as follows: Body Mass Index (BMI) > 27; psychiatric history or severe brain trauma; addiction, chronic medication affecting the central nervous system; smoking, excessive alcohol (> 14 units/week) or caffeine (> 3 cups/day) consumption; shift work in the past year; transmeridian travel in the past three months; moderate to severe subjective depression as measured by the Beck Depression Inventory (BDI) ^27^ (score > 19); poor sleep quality as assessed by the Pittsburgh Sleep Quality Index (PSQI) ^28^ (score > 7). Participants with sleep apnea (apnea hypopnea index > 15/hour; 2017 American Academy of Sleep Medicine criteria, version 2.4) were excluded based on an in-lab screening night of polysomnography. One participant, part of a twin pair, was excluded from the analyses so that the analysed sample included 363 participants (**Table 1**). Some EEGs were missing/lost/not recorded due to technical issues that were detected *a posteriori* for three to five participants per nights of sleep considered in this manuscript. No individual had missing EEGs for more than one night of sleep so that all 363 individuals contributed to at least part of the analyses reported here. The Epworth Sleepiness Scale ^29^ was used to characterize daytime sleepiness but was not used for inclusion. While most participants scored normal values (≤ 11), 28 participants had scores ranging from 12 to 15, corresponding to moderate daytime sleepiness. Because of an initial error in automatic evaluation of computerized questionnaires, seven participants had PSQI scores higher than cut-off (scores of 8 or 9). No participants were, however, taking sleep medication. To avoid reducing sensitivity, these participants were included in all analyses but removing them did not change statistical outcomes. Furthermore, IQ was estimated in all participants using the Raven Progressive Matrices ^30^. One item or more was not responded to by a few participants so that IQ was available in 347 participants. Likewise, the screening questionnaire did not include a clear question about number of years of educations, but was rather asking about current occupation, so that education was available in 300 participants. Including IQ or education in our statistical models (hence, in a reduced set of subject) did not affect the statistical outputs of the results presented below.

**Table 1.** Sample characteristics (mean ± SD).

|  |  |
| --- | --- |
| N | 363 |
| Sex | Men |
| Ethnicity | Caucasian |
| Age (y) | 22.10 ± 2.73 |
| Height (cm) | 180.39 ± 6.70 |
| Body mass index (BMI) (kg m<sup>-2</sup>) | 22.15 ± 2.31 |
| IQ* | 123.88 ± 11.14 |
| Education (y)** | 13.33 ± 1.60 |
| Mood | 3.00 ± 3.48 |
| Sleep quality | 3.46 ± 1.76 |
| Daytime sleepiness | 5.94 ± 3.54 |
| Chronotype | 50.11 ± 8.25 |
| Rest-activity Fragmentation (a.u.) | 0.10 ± 0.03 |
| Baseline sleep duration (min) | 451 ± 41 |

Mood was estimated by the 21-item Beck Depression Inventory II <sup>27</sup>, sleep quality by the Pittsburgh Sleep Quality Index (PSQI) <sup>28</sup>; daytime sleepiness by the Epworth Sleepiness Scale (ESS)<sup>29</sup>; chronotype by the Horne-Östberg questionnaire <sup>78</sup>. IQ was estimated using Raven Progressive Matrices <sup>30</sup>. Rest fragmentation (arbitrary units, a.u.) was estimated as the probability of transition from rest to activity during estimated sleep based on actigraphy data from the 3 weeks of imposed regular sleep <sup>11,44</sup>.
\* IQ was available for 347 participants. \*\* Number of years of education was available for 300 participants.

Although available in our laboratory, Ab- and tau-PET scans were not conducted in participants: it was felt to be unethical to expose them to an irradiation while results would most likely be normal.

### Experimental Protocol

Individual sleep-wake history was strictly controlled: during the three weeks preceding the in-lab experiment, participants were instructed to follow a regular sleep schedule according to their habitual sleep timing (+/-30 min for the first 2 weeks; +/-15 min for the last week). Actigraphy data showed that included participants faithfully followed the assigned schedules.

**Figure 1** provides an overview of the protocol. On Day 1, a urine drug test was performed (10-multipanel drug) before completing an adaptation night at habitual sleep/wake schedule during which a full polysomnography was recorded in order to screen for sleep related breathing disorders or periodic limb movements. On Day 2, participants left the lab with the instruction not to nap (checked with actigraphy). They returned to the laboratory at the end of Day 2, completed a baseline night of sleep under EEG monitoring at habitual sleep/wake schedule and remained in the laboratory until Day 7 under constant CCTV. A 12h sleep extension night under EEG and centered around habitual sleep mid-point was initiated on Day 3, in complete darkness with the instruction to try to sleep as much as possible. Day 4 included a 4h afternoon nap under EEG recording (centred 1h after the mid-point between morning wake-up time and evening sleep time) further dissipated any residual sleep need. What we termed the “before” night was also initiated on Day 4. It consisted in 8h sleep opportunity starting at habitual sleep time. During Day 5 and 6, participants remained awake for 40 hours under constant routine (CR) conditions [dim light < 5 lux, semi-recumbent position, 19°C ± 1, regular isocaloric food intake] before initiating a 12h recuperation night from habitual sleep time until 4h after habitual wake time. Except during sleep (darkness – 0 lux) and constant routine protocol (dim light < 5 lux), participants were maintained in normal room light levels oscillating between 50 and 1000 lux depending on location and gaze. Analyses of “before” night, nap and sleep deprivation protocol will be reported elsewhere. The current study focusses on baseline, extension and recovery nights of sleep.

**Figure 1:**
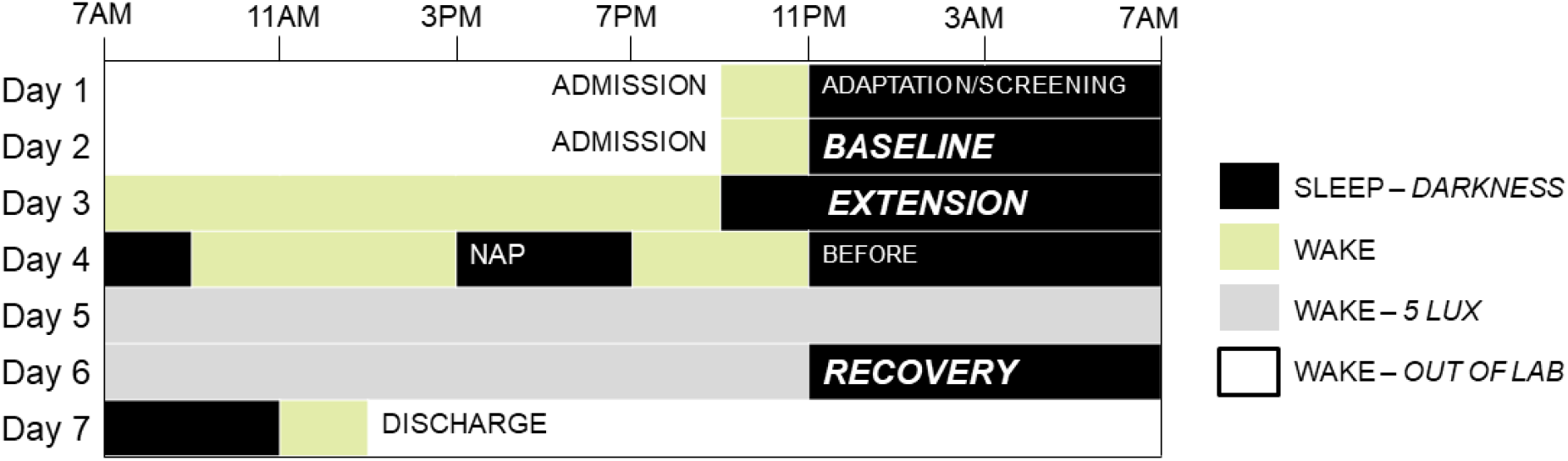
Overview of the protocol. Following 3 weeks of regular sleep at habitual times, 363 healthy young men aged ∼22 y complete a 7-day protocol (displayed for a participant sleeping from 11PM to 7PM). Adaptation/screening and baseline nights were scheduled at habitual sleep-wake times. Extension nights consisted of a 12h sleep opportunity centred around habitual sleep mid-point. Nap consisted of an afternoon 4h sleep opportunity. The “Before” (sleep deprivation) and recovery nights (from sleep deprivation) consisted of an 8h and 12h sleep opportunity respectively, all starting at habitual sleep time. Following the “before” night, volunteers completed a 40h sleep deprivation protocol under strictly controlled constant routine conditions in dim light. Sleep periods included in the current analyses are in bold and italic.

### EEG acquisitions and analyses

Sleep data were acquired using Vamp amplifiers (Brain Products, Germany). The electrode montage consisted of 10 EEG channels (F3, Fz, F4, C3, Cz, C4, Pz, O1, O2, A1; reference to right mastoid), 2 bipolar EOGs, 2 bipolar EMGs and 2 bipolar ECGs. Screening night of sleep also included respiration belts, oximeter and nasal flow, 2 electrodes on one leg, but included only Fz, C3, Cz, Pz, Oz and A1 channels. EEG data were re-referenced off-line to average mastoids. Scoring of sleep stages was performed automatically in 30-s epochs using a validated algorithm (ASEEGA, PHYSIP, Paris, France) ^31^ and according to 2017 American Academy of Sleep Medicine criteria, version 2.4. An automatic artefact detection algorithm with adapting thresholds ^32^ was further applied on scored data. Power spectrum was computed for each channel using a Fourier transform on successive 4-s bins, overlapping by 2-s., resulting in a 0.25 Hz frequency resolution. The night was divided into 30 min periods, from sleep onset until lights on. For each 30 min period, slow wave energy (SWE) was computed as the sum of generated power in the delta band (0.5 – 4 Hz range) during all the NREM 2 (N2) and NREM 3 (N3) epochs of the given period, after adjusting for the number of N2 and N3 epochs to account for artefacted data ^33^. As the frontal regions are most sensitive to sleep-wake history^34^, SWE was considered over the frontal electrodes (mean over F3, Fz, F4). To deal with the multiple comparison issue, we did not consider SWE over the other parts of the scalp ^35^. Additional analyses also considered cumulative power between 0.5 and 25 Hz during NREM and cumulative power between 2 to 6 Hz power during REM sleep as well through similar computation procedures.

### Genotyping and Imputation

Blood sample were collected on Day 2 for DNA analyses. The genotyping was performed using the Infinium OmniExpress-24 BeadChip (Illumina, San Diego, CA) based on Human Build 37 (GRCh37). Missingness of the SNP markers were below 20% in all individuals. Using PLINK software ^36^, we excluded the SNPs with a minor allele frequency (MAF) below 0.01, or Hardy-Weinberg disequilibrium (HWD) significance below 10^−4^. Markers with ambiguous alleles (A-T, T-A, G-C, C-G) were excluded as well. We finally ended with 511,729 SNPs. To investigate the relatedness between the individuals, using PLINK ╌genome command, we computed the identity by descent (IBD) estimates for all pairs of individuals. For 8 pairs, the composite pi-hat score was between 0.15 and 0.56 suggesting the existence of at least 3rd degree relatives in our cohort. We did not exclude any individuals at this level of analysis to keep the sample as large as possible, but removing one subject of each of these 8 pairs did not affect the statistical significance of any of the tests reported below. We merged our cohort with “1000 Genomes Project” ^37^ and employed principal component analyses (PCA) on the merged samples to see if our cohort was located in the European cluster (**Supplementary Figure S1A**). We further assess allele frequencies coherence of our cohort with the European subset of “1000 Genomes Project” (**Supplementary Figure S1B**). Markers with allele frequencies deviating more than 0.2 unit from European allele frequency were excluded (**Supplementary Figure S1C**). Genotype imputation was performed using “Sanger imputation server” by choosing “Haplotype Reference Consortium (release 1.1)” (HRC) as Reference Panel and the Pre-phasing algorithm EAGLE2. Post-imputation QC was then performed very similarly to the one of above (MAF < 0.01, HWD < 10^−4^, imputation quality score < 0.3). As a result of such filters, 7,554,592 variants remained for the analysis. However, to avoid having markers with allele frequencies deviating from European allele frequency, we computed the allele frequencies for the samples in our cohort after imputation and cross checked them with the European allele frequency (obtained from HRC Reference Consortium (release 1.1)) (**Supplementary Figure S1D**). The markers whose allele frequencies were deviating more than 0.2 unit from European allele frequency were excluded.

### Predicting Height

To validate common SNP assessments in our sample we predicted actual height based on Polygenic Scores computed based on a meta-analysis of a recent GWAS study ^38^ on around 700,000 individuals. We used all the variants in the meta-analysis that were included in our cohort [3121 SNPS out of 3290]. The procedure for calculating the Liability for height is the same as the one described in the following section. **Supplementary Figures S1E** visualize the Pearson correlation results between the actual values for Height and estimated genetic Liability of Height (r = 0.46, p = 10^−20^). Explained variance is very close to that reported previously ^38^, i.e. is 24.6%.

### Polygenic Risk Score (PRS)

Polygenic risk score (PRS) is defined as the sum of multiple single-nucleotide polymorphism alleles associated with the trait for an individual, weighted by the estimated effect sizes ^19,20^. We used the estimated effect sizes from a GWAS by Marioni et al. ^39^ which consisted of a meta-analyses of AD-by-proxy [UK Biobank data ^40^ - http://www.ukbiobank.ac.uk] and AD case-control data ^41^ for a total of 388,324 individuals (67,614 cases – 25,580 patients and 42,034 self-reported parental history of AD – and 320,710 controls). Marioni et al. reported that the genetic correlation between AD-by-proxy and AD case-control was very high and not significantly different from 1, so that the genetic associations they computed, and therefore the PRS we computed based on their summary statistics, were truly dealing with AD.

The best p-value threshold that should be applied to AD case-control summary statistics is not established yet. Previous studies employed very exclusive GWAS p-values (p∼10^−8^) ^42^ to more inclusive p-values (p = .5) ^25,43^, leading to the inclusion of effect sizes of a few tens to hundreds of thousands SNPs to compute AD PRS. Because we did not want to test all combinations of LD pruning and p-value thresholding, and then pick out the “best” one, we computed several PRS with different p-value thresholding and LD pruning combinations.

To generate a set of approximately independent SNPs in our sample, linkage disequlibrium (LD) clumping was performed using PLINK ^36^ on window size of 1000-kb using a pairwise r^2^ cut-off of 0.2 and a predetermined significance thresholds (*p*-value *<* 5 10^−8^, 10^−6^, 10^−4^, 0.001, 0.01, 0.05, 0.1, 0.3, 0.5, and 1). Due to the effect of APOE in chromosome 19, we used a more stringent criteria pairwise r^2^ cut-off of 0.01 for this chromosome. In addition, we also calculated the PRS using all the variants with no pruning, i.e. no correction for linkage disequilibrium, thereby selecting all SNPs for PRS construction. Although the later PRS was inevitably affected by complex LD structures, it was kept as one of the PRS. This procedure yielded 11 quantitative polygenic score, under each significance threshold, for each individual in our cohort.

### Height as a negative control

From the known and hypothesised biology, we did not expect any a priori association between the sleep phenotypes and a genetic liability for height. Therefore, we included an analysis of polygenic scores for height as a negative control, performing exactly the same association analyses as we did for liability to AD.

### Actigraphy data collection and analysis

Actigraphy data were collected with Actiwatch 4 devices (Cambridge Neurotechnology ltd, UK) worn on the non-dominant arm. Data consisted in the sum of activity counts over 60-second intervals. Data were analyzed with *pyActigraphy* (Version v0.1) ^44^ which implements the computation of state transition probabilities from rest to activity (kRA) ^11^. In order to better reflect sleep fragmentation, this probability was calculated only over sleep periods for each study’s participant. The sleep period is defined as the period comprised between the activity offset and onset times, derived from the average 24h activity profile. In addition, to mitigate the uncertainty on their exact timing, the offset and onset times were shifted by +15 min and −15min, respectively.

### Statistical Analysis

We employed general linear model (GLM) to test the associations between sleep metrics of interests as a dependent variable and the estimated PRS as an independent variables and age, BMI and TST as covariates. Prior to the analysis, we removed the outliers among the sleep metrics by excluding the samples lying beyond 4 times the standard deviation (the final number of individuals included in each analyses is reported below each dependent variable in the supplementary tables). All analyses were performed in Python.

In this study, we analysed multiple traits and multiple polygenic risk scores (PRS) for association. To control the experiment-wise false positive rate, we estimated the number of independent tests that we performed, and set an experiment-wise p-value threshold accordingly. Since the traits are phenotypically correlated with each other and the PRSs are also correlated, we used the correlation structure to estimate the equivalent number of tests, which is the number of independent tests that would result in the same overall observed variation.

For each correlation matrix of traits and PRS, we performed a singular value decomposition (SVD), ordered the resulting eigenvalues and calculated the sum of all eigenvalues. We then calculated the minimum number of linear combination of the traits that resulted in 99% of the variation. For the 5 EEG phenotypic sleep traits this estimate was 5, showing that they are not highly correlated. Likewise, for the 3 non-EEG phenotypic sleep traits this estimate was 3. For the 11 PRS for AD and height, the resulting number was 8 and 4, respectively, consistent with a higher correlation structure among the multiple height predictors. Therefore, our analyses with the 5 EEG sleep metrics implies a total number of 40 and 20 tests when confronted to AD-PRS and height-PRS respectively. Hence, for any of our trait-PRS combination to be statistically significant when taken multiple testing into account, the p-value threshold are 0.00125 and 0.0025 for AD and height, respectively. Similarly, our analyses with SWE in recovery and extension nights and with SWE rebound, each imply 8 tests and a p-value threshold of p = 0.00625, while our analyses with 3 non-EEG sleep metrics 24 tests and a p-value threshold of p = 0.0021. Additional analyses compared lower and higher PRS quartile (i.e. 90 individuals with lowest AD PRS and 90 individuals with highest PRS) as well as APOE ε4 carriers vs. non carriers. For these analyses, groups were compared through t-tests.

We compute the minimum detectable effect size given our sample size. According to G-Power 3 (version 3.1.9.4) ^45^, taking into account a power of .8, an error rate α of 0.00625 (cf. above), with a sample size of 363, we were in a position to detect medium effect sizes r > 0.19 [confidence interval: 0.09-0.29] within a linear multiple regression framework including 7 predictors.

## Results

### Polygenic risk for AD is associated with the generation of slow waves during sleep

PRS were computed as the weighted sum of the effect sizes of the AD-associated SNPs, obtained from summary statistics of AD cases vs. controls GWAS ^19,20^. PRS can indicate the presence of a genetic signal in moderate sample size studies ^19,23^ as long as it is computed based on a very large GWAS ^46,47^. We therefore used the summary statistics of one of the largest AD-GWAS available to date (N = 388,324) ^39^ to compute individual PRS for AD in our sample and related these to sleep EEG characteristics following multiple quality control steps (cf. **Supplementary Figure S1**).

We first focused on baseline sleep, as it is most representative of habitual sleep, to evaluate sleep metrics that might be associated with AD liability. Given our sample size, we reduced the multiple comparison burden by selecting *a priori* variables of interest among electrophysiology sleep metrics that have previously been related to Aβ and Tau in cognitively normal older adults: sleep onset latency [SOL] ^9,48^, duration of wakefulness after sleep onset [WASO] ^48^, duration of REM sleep ^10^, slow wave energy [SWE] during NREM sleep ^7,8^, i.e. the cumulated power in the 0.5-4 Hz EEG band, and hourly rate of micro-arousals during sleep ^14^. To compute PRS, one considers SNPs below a p-value threshold in the reference GWAS; the optimal threshold for SNP selection to best compute a PRS for AD is not established. To avoid bias in the threshold selection, we opted for computing PRS based on increasingly inclusive p-value thresholds (including SNPs reaching GWAS significance – p < 5×10^−8^ - to very liberal p < 1), whilst also pruning SNPs based on their correlation structure (i.e. linkage disequilibrium) (**Supplementary Table S1**) ^25,43^. In addition, we performed a PRS analysis using all SNPs without any selection.

General linear model (GLM) analyses controlling for age, body mass index (BMI) and total sleep time (TST), revealed an significant association between baseline night SWE and AD PRS (p < 0.02; **β ≥ 0**.**12**) from a p-value threshold of p=0.05 up to selecting all SNPs; the association reached stringent experiment-wise correction for multiple comparisons when computing PRS using all SNPs, i.e. with potential linkage disequilibrium bias (see methods; **β = 0**.**17**; **Figure 2A; Supplementary Table S2**). We performed a negative control analysis using a PRS for height, a variable for which no association with sleep metrics was expected, and found no association (**Supplementary Figure S2A**). The association between AD PRS and SWE was positive (**Figure 2B**) indicating that higher SWE was associated with higher AD-PRS. SWE was also positively associated with TST (**Supplementary Table S2**), which was expected since TST conditions the opportunity to generate slow waves, and negatively with age, which is in line with the literature ^49^ but may still be surprising given the young age of our sample. Importantly, since GLM included TST and age, they are not driving the association we find between SWE and PRS for AD. Furthermore, we performed two additional analyses seeking for associations between PRS for AD and IQ or education, variable for which negative associations with AD pathophysiology were previously reported ^50^, and found no associations (**Supplementary Figure S2B**). The link between PRS for AD and SWE may therefore more consistent (i.e. less variable) than the link between AD and IQ or education.

**Figure 2:**
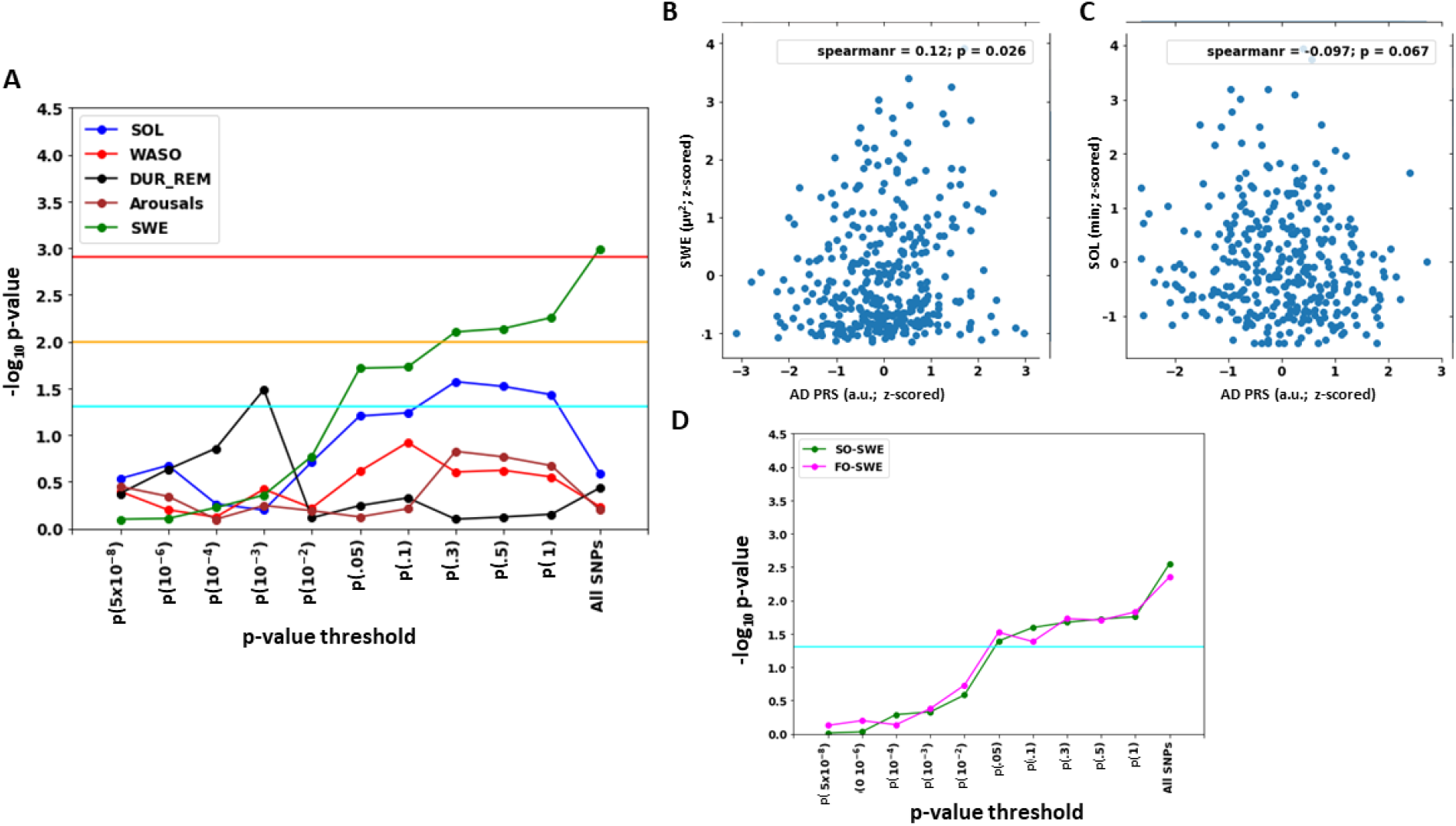
Associations between Polygenic Risk Score (PRS) for AD and baseline night sleep metrics. **A.** Statistical outcomes of GLMs with five sleep metrics of interest vs. AD PRS from conservative (p < 5×10^−8^) p-value threshold to using all SNPs (N=356). GLMs are corrected for age, BMI and total sleep time (TST). Negative log transformation of p-values of the associations are presented on the vertical axis. Horizontal lines in A and D indicate different p-values thresholds: light blue = .05 (uncorrected); orange= .01 (corrected for 5 sleep metrics); red = 0.00125 (experiment-wise correction; see methods). SOL: sleep onset latency; WASO: wake time after sleep onset; DUR_REM: duration of REM sleep; arousal: hourly rate of micro-arousal during sleep; SWE: slow wave energy in NREM sleep (0.5-4Hz) **B.** Positive association between SWE during baseline night and AD PRS including All SNPs (N=356). Spearman correlation r is reported for completeness (r = .12, p = .02), refer to main text Table S2 for statistical outputs of GLMs. **C.** Negative association between SOL during baseline night and AD PRS for p < 0.3. Spearman correlation r is reported for completeness (r = −.11, p = .03), refer to main text Table S2 for statistical outputs of GLMs (N=356). **D.** GLMs including SWE separated in the slower (SO-SWE; 0.5-1Hz) and faster (FO-SWE; 1.25-4Hz) frequency range from conservative p-value thresholds to using all SNPs (N=356). Horizontal blue line indicate p = 0.05 significance level. GLMs are corrected for age, BMI and TST. Refer to main text Table S3 for statistical outputs of GLMs.

Sleep onset latency (SOL) also reached significant association with AD PRS from a p-value threshold of p=0.05 up to p = 1 (p ≤ 0.04; **β = −0**.**11**), but significance did not reach stringent experiment-wise correction for multiple comparisons (**Figure 2A; Supplementary Table S2**). Hence, this result has to be considered with caution and will not be extensively commented upon. It is interesting to note, however, that the association between PRS for AD and SOL is negative, with higher PRS associating with shorter sleep latency (**Figure 2C**). Of note, REM% reached uncorrected significance (p < 0.05) for thresholding at p=0.05 (**β = 0**.**1**), with a positive association with AD PRS (**Figure 2A; Supplementary Table S2**), but, since it is observed for only one p-value threshold, this will not be discussed any further.

These results indicate that, particularly when considering all SNPs to construct the AD PRS, the overnight power of the slow waves generated during Non-REM sleep, which is a widely accepted measure of sleep need ^51^, is linearly and positively associated with AD genetic liability. This finding suggests that individuals with a higher genetic liability for AD have a higher need for sleep. This idea is further reinforced by the fact that association between SWE and AD PRS is also significant when only considering SWE of the first hour of sleep ^51^ (**Supplementary Figure S3 & Table S3**), and the potential negative association with SOL, which depends in part on sleep need.

Since slow oscillations (SO), i.e. EEG slow waves < 1 Hz, may be distinct from faster slow waves ^52^, we further decomposed SWE into SO-SWE (0.5-1Hz) and faster-oscillations—SWE (FO-SWE; 1.25 – 4 Hz). Both SO-SWE and FO-SWE were similarly and significantly associated with AD PRS and for the same p-value thresholds (**Figure 2D; Supplementary Table S3**). The association we found between SWE and AD PRS does not appear therefore to arise exclusively from either slower or faster slow waves.

### Recovery sleep, slow wave sleep rebound and extension night

When considering sleep EEG of the other nights, we only included SWE, as it is the only sleep metric that was associated with PRS for AD at stringent correction for multiple comparisons threshold. Similarly to baseline night, when considering SWE during the recovery night that followed total sleep deprivation, GLM including age, BMI and TST, reveal that SWE and AD PRS are significantly associated (p ≤ 0.04; **β ≥ 0**.**11**) from p-value thresholding at p=0.1 up to using all SNPs (**Figure 3A; Supplementary Table S4**), and the association reached stringent experiment-wise correction for multiple comparisons at p-value threshold of p=1. Again, the association was positive with higher SWE associated with higher AD PRS (**Figure 3B**) and results were similar when considering only SWE of the first hour of sleep (**Supplementary Figure S3 & Table S4**). Individuals typically produce more sleep slow waves in response to sleep loss, as part of the homeostatic regulation of sleep ^53^. Therefore, individuals with higher need for sleep after sleep loss have a high PRS for AD.

**Figure 3:**
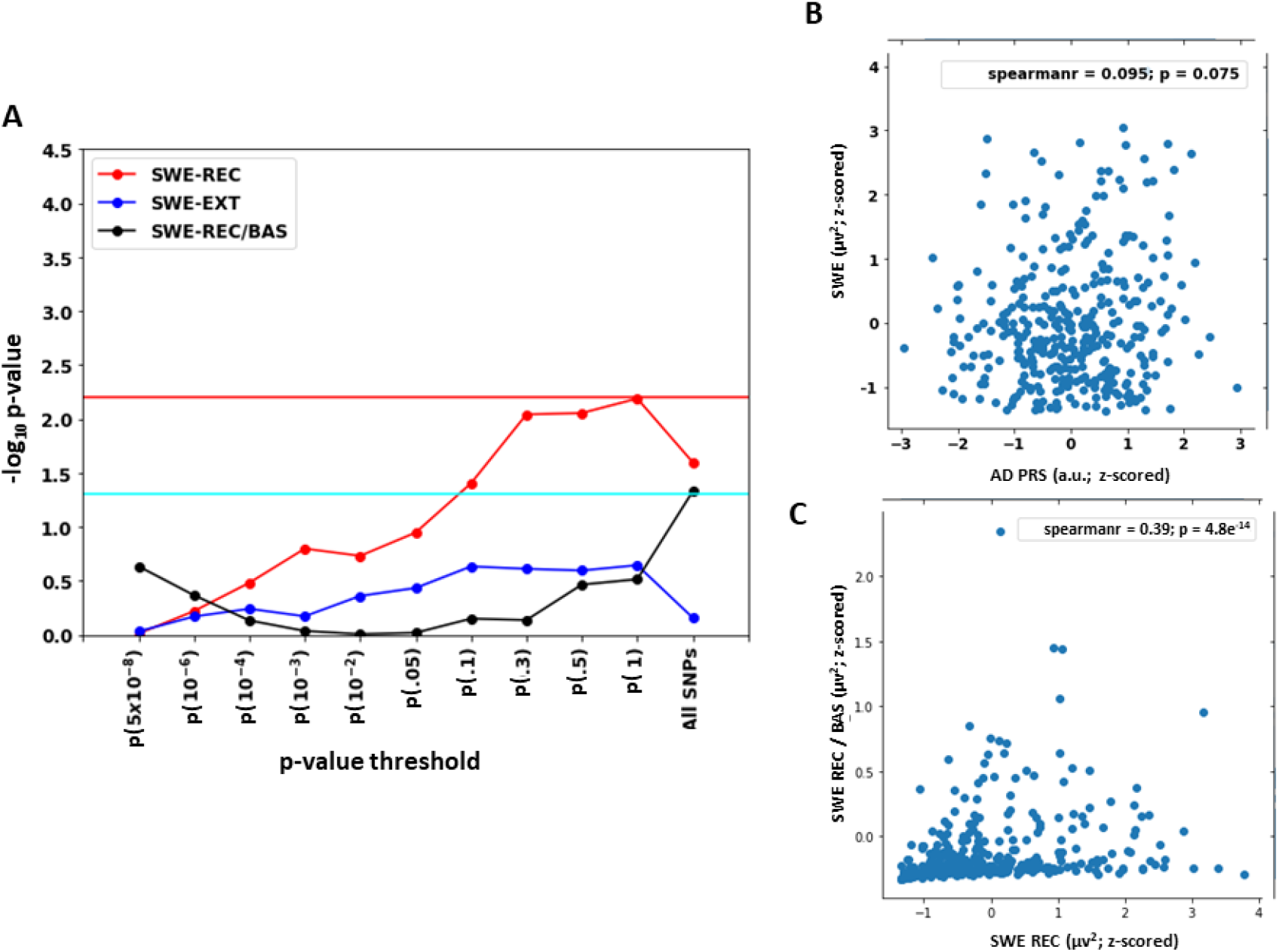
Associations between Polygenic Risk Score (PRS) for AD and slow wave energy (SWE) during recovery and extension nights and with SWE rebound. **A.** Statistical outcomes of GLMs with SWE (0.5-4Hz) in the recovery (REC; N=353) and extension (EXT; N=356) nights and with SWE rebound (REC/BAS; N=344) vs. AD PRS from conservative (p < 5×10^−8^) to inclusive (p < 1) p-value level and using all SNPs. SWE rebound consist in the ratio between SWE in the first hour of sleep of recovery and baseline nights. GLMs are corrected for age and BMI, and TST for REC and EXT. Negative log transformation of p-values of the associations are presented on the vertical axis. Horizontal lines indicate different p-values thresholds: light blue = .05 (uncorrected); red = 0.00625 (experiment-wise correction; see methods). **B.** Positive association between SWE during recovery night and AD PRS at p < 1. Spearman correlation r is reported for completeness (r = .01, p = .06), srefer to main text Table S4 for statistical outputs of GLMs (353). **C.** Positive association between SWE during recovery and SWE rebound (SWE REC/BAS): Spearman correlation r = .36, p < .001 (N=344).

Slow wave sleep rebound quantifies the physiological response to a lack of sleep based on the relative changes from normal sleep to recovery sleep following sleep loss. We computed the ratio between the initial SWE (1h of sleep) during recuperation and baseline nights to assess SWE rebound. GLM analysis, including age and BMI, indicated that SWE rebound reached significant association with AD PRS when including all SNPs (**β = −0**.**11**), but significance did not reach stringent experiment-wise correction for multiple comparisons (**Figure 3A; Supplementary Table S4**). Sleep rebound is driven by sleep homeostasis which tightly regulates sleep duration and intensity based on prior sleep-wake history ^35^. Since we observe an association with AD PRS for a single p-value threshold at uncorrected p-value our findings suggest that, in our sample, AD PRS was not tightly associated with sleep homeostatic response. Interestingly though, Spearman’s correlation indicated that SWE rebound was correlated to SWE during the recovery night (r = 0.39, p <10^−14^; **Figure 3C)**.

We then considered SWE during the extension night and PRS for AD in a GLM, including age, BMI and TST. Results indicated that extension night SWE was not significantly linked to AD PRS. This may be because sleep timing for this particular night affects sleep quality ^35,51^ (**Figure 3A**). In contrast to baseline and recuperation sleep periods which were initiated at habitual sleep time, sleep extension started 2 hours before habitual sleep time, covering the end of a period known as the evening “wake-maintenance zone” corresponding to the time at which the circadian system maximally promotes wakefulness ^51^. In addition, the circadian system is known to affect the relative content in Non-REM and REM sleep as well as in different EEG frequencies ^35,51^. Therefore, the imposed 2h advance of sleep time during the extension night affected sleep quality, which may have reduced the association between SWE and AD PRS found with baseline and recovery nights.

### Polygenic risk for AD is associated with increased subjective daytime sleepiness

We next focused on the non-EEG sleep metrics of our protocol and explored their potential association with AD PRS. Based on the 3 weeks of actigraphy with imposed regular habitual sleep time at home, we computed the probability of transition from rest to activity during the sleep period [kRA; ^11^]. kRA is a proxy for sleep fragmentation and has been associated with cognitive decline and the risk for developing AD in cognitively normal older adults [mean age 81.6 y ^11^]. kRA showed a negative association (higher AD PRS is associated with less fragmented sleep) with PRS for AD for two p-value thresholds, p=5 × 10^−8^ and p = 10^−8^ (**Figure 4A; Supplementary Table S5**), but did not reach stringent experiment-wise correction for multiple comparisons (p < 0.002); it will not be further discussed.

**Figure 4:**
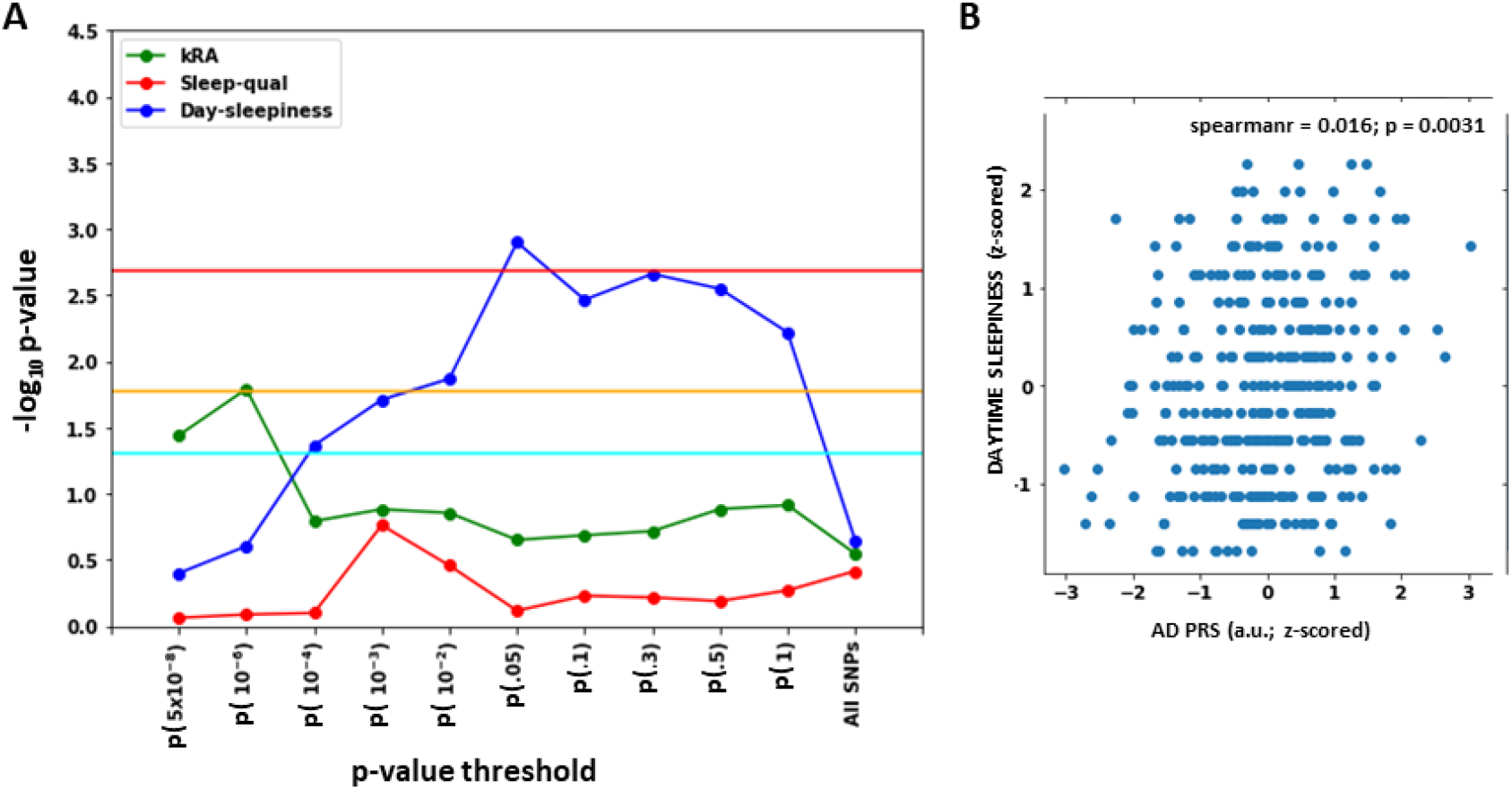
Associations between Polygenic Risk Score (PRS) for AD and non-EEG sleep metrics. **A.** Statistical outcomes of GLMs with actimetry-assessed sleep fragmentation (kRA; N=361), subjective sleep quality (Sleep-qual; N=363) and subjective daytime sleepiness (Day-sleepiness; N=363) vs. AD PRS from conservative (p < 5×10^−8^) to inclusive (p < 1) p-value thresholds and using all SNPs. GLMs are corrected for age and BMI. Negative log transformation of p-values of the associations are presented on the vertical axis. Horizontal lines indicate different p-values: light blue = .05 (uncorrected); orange= .016 (corrected for 3 sleep metrics); red = 0.002 (experiment-wise correction). **B.** Positive association between subjective daytime sleepiness and AD PRS at p < 0.05 (N=363). Linear regression line shown for display purposes only; refer to main text and Table S5 for statistical outputs of GLMs.

Two questionnaires assessed habitual subjective sleep quality and daytime sleepiness before the start of the protocol. Subjective sleep quality was not significantly associated with AD PRS. By contrast, subjective daytime sleepiness was significantly associated with PRS for AD (p < 0.05; **β ≥ 0**.**11**) from thresholding at p < 10^−4^ up to a threshold of p < 1 and at stringent experiment-wise correction for multiple comparisons at p-value thresholds of p < 0.05 and p < 0.3 (**β ≥ 0**.**16**; **Figure 4A; Supplementary Table S5**). The association was positive indicating that higher habitual subjective daytime sleepiness was associated with higher AD PRS (**Figure 4B**). This shows that the association between AD PRS and sleep need, as assessed by electrophysiology, is not a mere effect of the protocol and is mirrored at the behavioural level during habitual daytime functioning (outside the experimental protocol). Importantly the vast majority of participants had no or mild levels of sleepiness with a minority (N = 28) reporting moderate level of daytime sleepiness; the association with daytime sleepiness is therefore not driven by extreme or clinically relevant sleepiness levels but rather by ordinary variability in healthy young individuals.

## Discussion

We provide evidence that genetic liability for AD is related to sleep characteristics and daytime sleepiness in young adults (aged 18 to 31 y), i.e. decades before typical onset age of clinical AD symptoms and at an age at which current AD biomarkers are typically negative. Our sample size is modest for the detection of small effect size associations, and we do not include a replication sample, so the present results should be considered as a proof-of-concept for linking AD liability and sleep in young adults. We emphasize, however, that the unique deep phenotyping of our protocol in hundreds of participants, based on gold standard electrophysiology and comprising different sleep conditions, one the one hand, makes the creation of a replication sample difficult but, on the other, undoubtedly increased the sensitivity of our analyses so that we could find associations that survived stringent correction for multiple comparisons. In addition, we performed a negative control analysis using a PRS for height variables for which no association with sleep metrics was expected, and found no association. Furthermore the absence of links between PRS for AD and IQ and education suggest that the association between PRS for AD and SWE is more stable across subjects than the link previously isolate between AD and IQ or education ^50^. Importantly, our protocol provides links between disease risk and sleep physiology in contrast to coarser phenotyping based on sleep questionnaires or actimetry alone. Furthermore, to increase the genetic uniformity of the sample, we only included Caucasian men within a narrow age range; they were healthy and devoid of any sleep disorders or sleep complaints and their prior sleep-wake history was recorded and stable. In this carefully selected homogenous sample, we show that higher PRS for AD was associated with producing denser or larger slow waves during baseline and recovery night time sleep, potentially with large slow wave sleep rebound following sleep deprivation, and with reporting higher daytime sleepiness.

Larger and more abundant slow waves during habitual sleep in young and healthy individuals can result from an increased sleep need due to insufficient prior sleep ^53^. This appears unlikely: prior sleep-wake history was stringently controlled for 3 weeks prior to entering the lab, ruling out undue sleep deprivation, sleep restriction or disrupted rhythmicity. Moreover, throughout the protocol, participants followed their own sleep schedule, a regime that should not expose them to important chronic sleep restriction. Finally, SWE during the sleep extension night did not significantly correlate with subjective daytime sleepiness (Spearman’s correlation r = 0.08, p = 0.11), supporting the idea that, when given a longer sleep opportunity, individuals with higher and yet normal daytime sleepiness did not sleep more intensely to recover a putative prior sleep debt. Alternatively, increased slow wave density and/or intensity could reflect a faster build-up of sleep need ^54^. Indeed, sleep homeostasis is thought to result from molecular and cellular changes induced by waking brain function and behaviour ^55,56^. Synaptic potentiation and increased synaptic strength resulting from waking experience are reflected in a progressive increased cortical excitability during wakefulness ^57,58^ and an increase in slow wave activity during subsequent sleep ^55,56^. Likewise, extracellular glutamate concentration and glutamatergic receptor density increase with time awake and affect brain function ^59,60^. Here, SWE rebound following sleep loss, i.e. the ratio between baseline and recovery sleep, was only significantly associated with high PRS for AD for one p-value threshold and at uncorrected significance threshold, but was strongly associated with SWE during recovery sleep. We therefore find only partial evidence for this second hypothesis, which will require more investigations.

How are these findings related to AD? The answer to this question remains speculative because the time course of AD processes across lifespan is still poorly understood. In transgenic mice, neuronal activity locally increases the level of Aβ in the interstitial fluid and drives local Aβ aggregation ^61^. The progressive Aβ deposition ultimately disrupts local functional connectivity and increases regional vulnerability to subsequent Aβ deposition ^62^. We might thus hypothesize that individuals with more intense brain activity during wakefulness (and therefore also during sleep) would also be exposed to larger Aβ extracellular levels and a greater risk of developing Aβ deposits. This hypothesis appears unlikely for the following reasons. First, post mortem examinations show that the earliest evidence of Aβ deposits (stage 1 ^63^) is not observed before 30 y ^64^. Second, Aβ oligomers might be released and exert their detrimental effect on brain function at an earlier age. However, in transgenic mice, sleep-wakefulness cycle and diurnal fluctuation in brain extracellular Aβ remain normal until plaque formation ^65^.

By contrast, given the age range of our population sample, the reported topography of pretangles at this age ^16^ and the power of PRS for AD to discriminate AD patients in case-control samples ^43^, higher PRS in our young sample might reflect the influence of incipient Tau aggregation onto sleep regulation through the LC (and other non-thalamic cortically-projecting nuclei, as raphe nuclei) ^16^. Tau, an intracellular protein, is also detected in the extracellular space. Over and above a low level constitutive tau secretion ^66^, neuronal activity increases the release of tau in the extracellular space ^67^, thereby participating in enhancing tau spread and tau pathology in vivo ^68^. Moreover, early electrophysiological changes indicative of hyperexcitability are observed in intact neurons from transgenic tau mice ^69^. In the cerebral cortex of tau transgenic mice, glutamatergic and GABAergic neurons are in a hypermetabolic state, characterized by a relative increase in production of glutamate ^70^. By contrast, decreasing tau in epilepsy-prone transgenic mice reduces neuronal hyperexcitability ^71^. These findings would suggest that a strong cerebral activity during wakefulness would result in a higher daily average in perceived sleepiness, a substantial tau release – which lead to the formation of pretangle aggregates - and an enhanced sleep homeostasis processes, as indicated by denser and larger slow waves.

The reasons for the vulnerability of LC to Tau aggregation are not established but might reside in its constant recruitment for essential functions, its energy demanding and ubiquitous brain connections, its high vascularization or its higher susceptibility to oxidative stress ^15^. Although it tantalizing to hypothesize that tau pretangle aggregates are involved in the mechanisms linking slow wave sleep and AD liability, one can also speculate that it is the LC intrinsic characteristics that are related to tau vulnerability (subsequent) that associated with PRS for AD, meaning that the association would not necessarily require the presence of tau to be detected.

On the other hand, in tau transgenic mice, misfolded and hyperphosphorylated tau alters hippocampal synaptic plasticity ^72^, eventually induces a loss of hippocampal LTP and causes reduction of synaptic proteins and dendritic spines ^73 74^. These findings would predict a lower sleep need in participants with high AD liability. However, it is possible that these detrimental processes take place later on in the development of the disease or emerge from an interaction between tau and Ab ^75 76^. Accordingly, in older adults, significant associations, opposite to the current findings, were observed between slow wave sleep and risk for AD based on PET biomarkers ^7,8^: higher Aβ ^7^ or tau NFT ^8^ burdens were associated with lower sleep slow wave EEG power. Our results suggest therefore that the association between AD risk and sleep homeostasis changes with age: at an early stage, dense and large slow waves would be associated with increased AD risk. Later on, the ability to generate slow waves would play a protective role against AD risk. Deep sleep phenotyping across all ages and/or in long term longitudinal studies will have to test this hypothesis.

We emphasize that the cross sectional nature of our study, precludes any causal interpretation of the association we find between AD and sleep. We further stress that PRS estimation based on all available SNPs may be biased by complex linkage disequilibrium (LD) between SNPs. Since we find similar p-values when pruning SNPs for LD at other p-value thresholds, we are confident that the likely bias is not the main driver of the effects we report. Furthermore, our sample only include men and cannot therefore be extended to the entire population. Women have been reported to have different sleep characteristics, including the production of more numerous and intense slow waves during sleep ^77^. It is also worth mentioning that we cannot isolate in our findings the specific contributions of the circadian timing system, which is the second fundamental mechanism regulating sleep and wakefulness ^35^. Although we find significant association between AD PRS and baseline/recovery SWE and daytime sleepiness across similar p-value thresholds, more research is also required to determine how many SNPs one has to include, i.e. what SNP selection strategy should be used to best predict AD. Previous studies support that using a lenient p-value thresholds is successful in doing so ^25,43^, thus we are confident that our finding are related to AD liability. Our PRS calculation was stringently controlled for the weight of chromosome 19 (see methods) to avoid excessive contribution from Apolipoprotein E (APOE) genotype, which is the genetic trait most associated with sporadic AD. When comparing APOE ε4 carriers genotype vs. non-carriers, no significant difference in baseline night SWE and daytime sleepiness was observed **(Supplementary Figure S4)**, in line with our findings that a large number of SNPs is required to find an association between SWE and PRS for AD.

The specificity of our findings for a given EEG frequency band and/or for NREM remains to be fully established. As many previous studies on linking sleep and AD risk [e.g. ^7,8^], we only focussed on a limited set of sleep metrics, and included a single power measure over a given frequency band. Although not the focus of the present paper, we computed SWE, relative SWE (i.e. ratio between SWE and overnight total NREM power), overnight cumulated total power during NREM sleep and overnight cumulated power in the 2 to 6 Hz band during REM sleep of the baseline night in individuals among the higher and lower AD PRS quartile **(Supplementary Figure S5)**. This simple analyses indicates that individuals with 25% highest AD PRS had higher power than individuals with 25% lowest AD PRS for all three absolute measures (t-test; p ≤ 0.01 but not for relative SWE (p = 0.14), suggesting that our findings may not be specific to NREM sleep and SWE. We emphasize, however, that, given our modest sample size, our analyses was not planned to address such question. This first preliminary analysis warrants future studies with larger sample size ensuring sufficient power when using a larger set of sleep metrics. Since we also find that daytime sleepiness, a wakefulness trait, is associated with PRS for AD, and because of the link between tau protein and cortical excitability ^71^, neuronal activity synchrony during wakefulness should be associated with the risk for developing AD to assess whether isolated links are specific to sleep.

In conclusion, we find that denser and/or more intense sleep slow waves during baseline and recovery sleep and daytime sleepiness are associated with the genetic liability for AD in young and healthy young men. This finding supports that sleep slow wave and sleepiness measures may help early detection of an increased risk for AD and reinforce the idea that sleep may be an efficient intervention target for AD. Similarly to most studies associating PRS to phenotypes of interest [e.g. ^23–25,46^], the effects we isolated constitute relatively small effects (r < 0.2), however, recalling that sleep must be envisaged within the multifactorial aspect of a complex disease such as AD ^4^.

## Data Availability

The authors declare that the data supporting the findings of this study are available from
the corresponding author upon request.

## Author contributions

Study concept and design: V.M., S.N.A., M.G., D.J.D., P.M., P.M.V. and G.V. Data acquisition: V.M., M.J., C.Me. Data analyses: V.M., E.K., P.G., C.B., M.B., W.C., N.A. and G.V. Interpretation: V.M., E.K., P.G., D.J.D., P.M., P.M.V., G.V. Analyses support: E.S., A.L., C.P., L.Y., E.B., M.E., M.V., D.C., C.Mo., G.H., C.D., F.C., M.G., C.S. Manuscript draft: V.M., E.K., P.G., D.J.D., P.M., P.M.V., G.V. Revised manuscript: all authors.

### Acknowledgments

We thank E. Balteau, C. Borsu, N. Cai, A. Claes, G. Gaggioni, A. Gobalek, B. Herbillon, P. Hawotte, E. Lambot, B. Lauricella, M. Lennertz, J.Q.L. Ly, M. Micho, G. Salmon and A. Shaffii-Le Bourdiec for their help in different steps of the study.

P.G., M.V., C.S., F.C., C.P. and G.V. are supported by the Fonds de la Recherche Scientifique - FNRS-Belgium. The study was supported by the Wallonia-Brussels Federation (Actions de Recherche Concertées - ARC - 09/14-03), WELBIO/Walloon Excellence in Life Sciences and Biotechnology Grant (WELBIO-CR-2010-06E), FNRS-Belgium (FRS-FNRS, F.4513.17 & T.0242.19 & 3.4516.11), University of Liège (ULiège), Fondation Simone et Pierre Clerdent, European Regional Development Fund (Radiomed project), Fonds Léon Fredericq. DJD is supported by the UK Dementia Research Institute (DRI).

## Preprint repository

This manuscript is available on the preprint server medRxiv (https://www.medrxiv.org/content/10.1101/2020.02.26.20027912v1).

## ONLINE SUPPLEMENTARY INFORMATION

**Figure S1:**
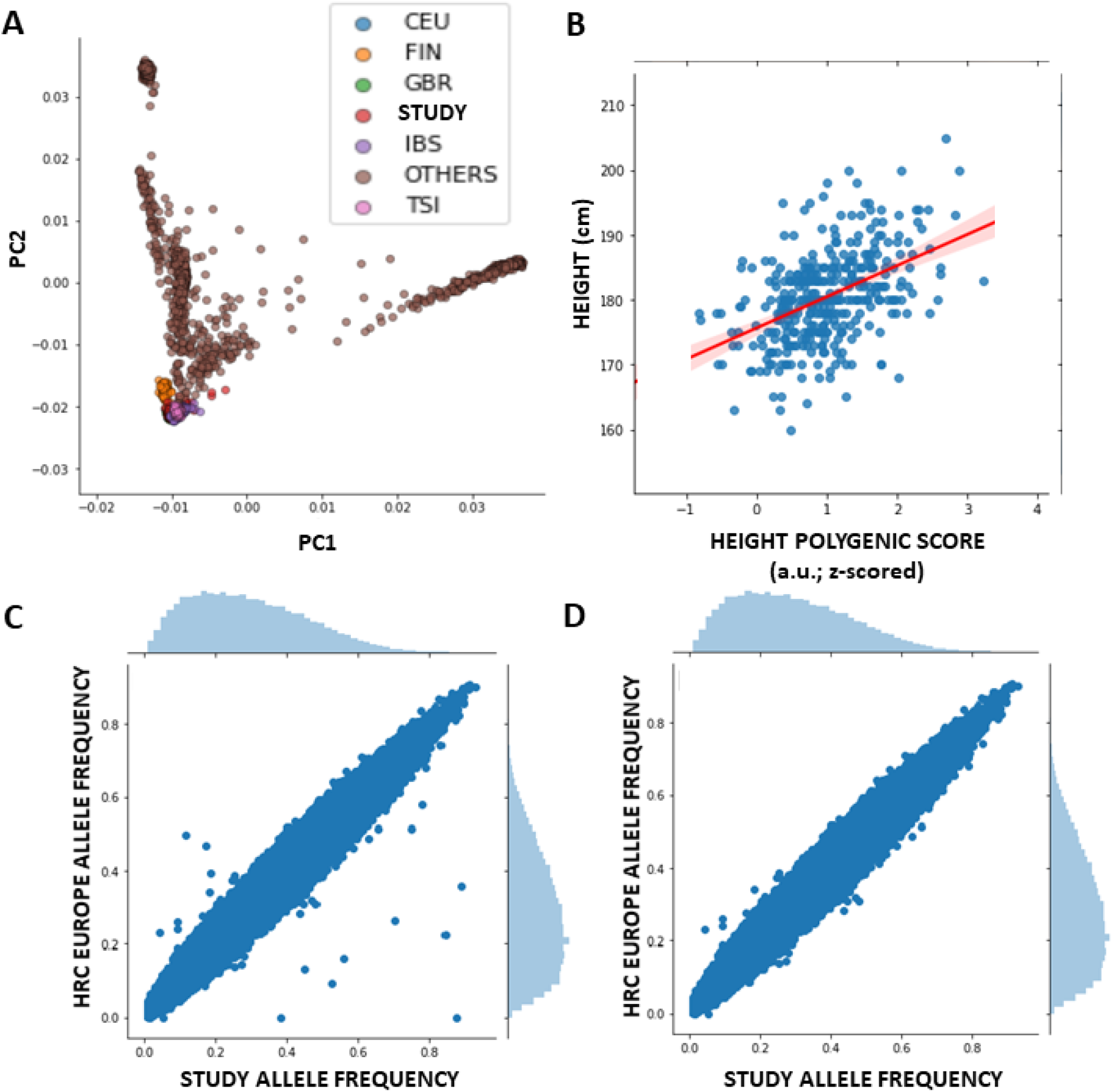
quality checks (QC) performed during genetic data processing. **A.** Principal component (PC) analysis of our data (after merging with genome 1k). PC1 vs. PC2 is displayed for our European (Caucasian) study sample (STUDY) and for other ethnicities provided by Haplotype Reference Consortium (HRC) Europe release 1.1. CEU: Utah Residents (CEPH) with Northern and Western European Ancestry; FIN: Finnish in Finland; GBR: British in England and Scotland; IBS: Iberia in Spain; TSI: Toscani in Italia. Our study sample clusters at the same position as Europeans. **B.** To validate common SNP assessments in our sample, we predicted height of our volunteers based on a meta-analysis of height GWAS studies ^75^. Predicted Liability for Height was significantly associated with the actual values in our sample (r = 0.46; p < 10^−20^) with similar value as reported in Yengo et al. (2018) (r = 0.49). **C-D**. Allele frequencies in our sample were compared to HRC Europe reference data and SNP deviating more than 0.2 unit from European allele frequency were excluded [**C**. Original data. **D**. Data after removal of deviant allele].

**Figure S2:**
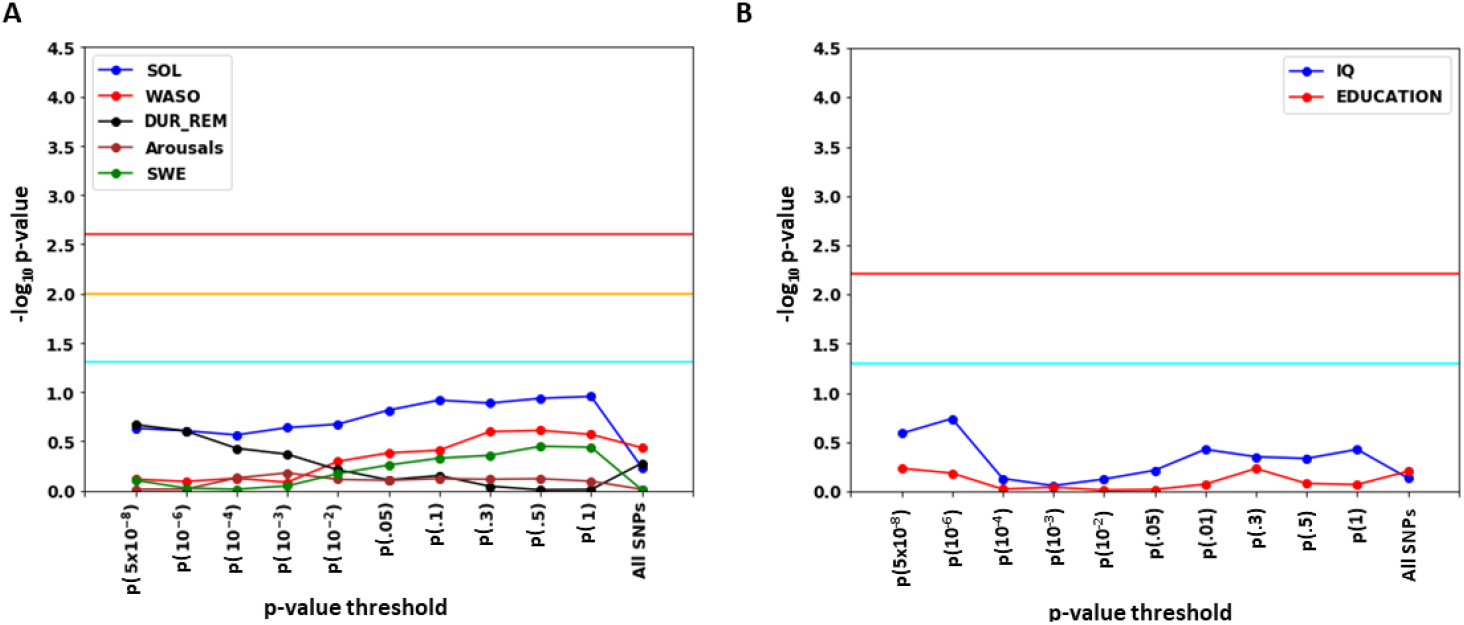
Negative controls analysis and correlation between AD PRS and IQ or education. **A.** Statistical outcomes of GLMs with five sleep metrics of interest vs. polygenic prediction of individual height from conservative (p < 5×10^−8^) p-value threshold to using all SNPs. Negative log transformation of p-values of the associations are presented on the vertical axis. Horizontal lines indicated different p-values thresholds: light blue = .05 (uncorrected); orange= .01 (corrected for 5 sleep metrics); red = 0.0025 (experiment-wise correction). As expected height prediction is not associated with any of the sleep metrics further validating our main finding. **B.** Statistical outcomes of GLMs with IQ (N=347) and number of years of education (N=300) vs. PRS for AD from conservative (p < 5×10^−8^) p-value threshold to using all SNPs. Negative log transformation of p-values of the associations are presented on the vertical axis. Horizontal lines indicated different p-value thresholds: light blue = .05 (uncorrected); red = 0.00625 (correction for 8 independent PRS). As expected IQ and education are not associated with PRS for AD.

**Figure S3:**
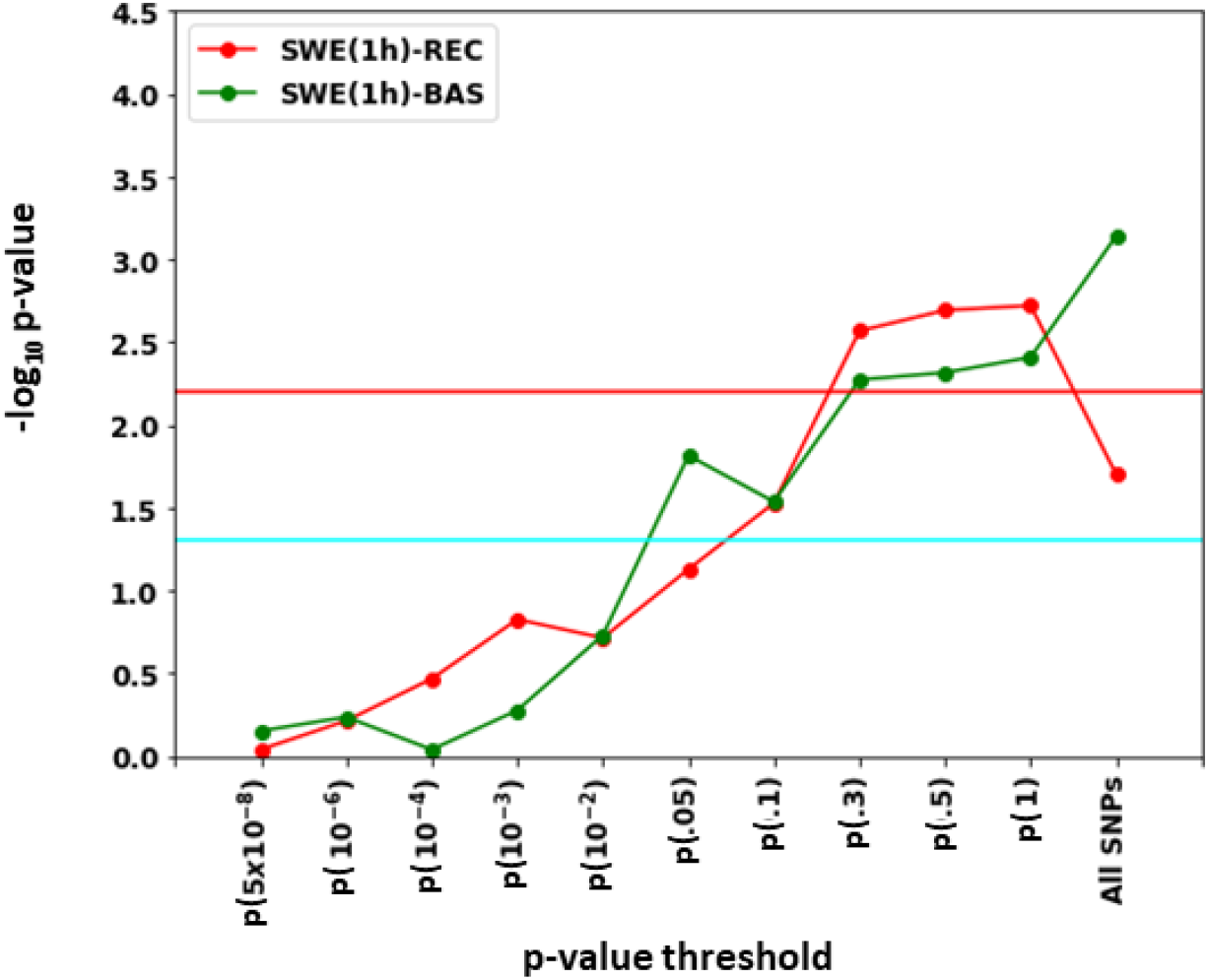
Associations between Polygenic Risk Score (PRS) for AD and SWE in the first hour of sleep for baseline (BAS), recovery (REC) and extension (EXT) nights. Statistical outcomes of GLMs with SWE (0.5-4Hz) of each night type in the first hour of sleep (SWE(1h)) vs. AD PRS from conservative (p < 5×10^−8^) p-value threshold to using all SNPs. GLMs are corrected for age and BMI. Negative log transformation of p-values of the associations are presented on the vertical axis. Horizontal lines indicate different p-values thresholds: light blue = .05 (uncorrected); red = 0.00625 (experiment-wise correction; see methods).

**Figure S4:**
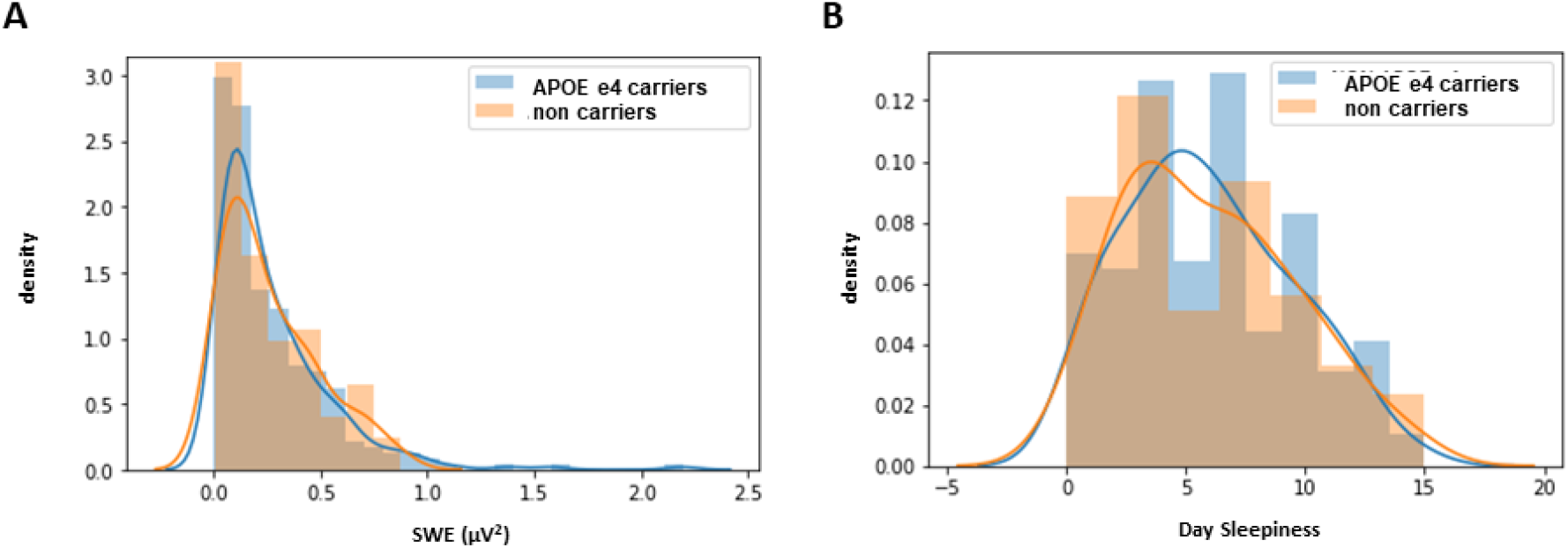
No difference in SWE and daytime sleepiness between APOE e4 carriers and non-carriers. Distribution of APOE e4 Carriers and Non Carriers for SWE during baseline night (**A**) and daytime sleepiness (**B**). T-tests between APOE e4 carriers (N = 100) and Non carriers (N = 263) show no significant difference in case of SWE (p = 0.84) and daytime sleepiness (p = 0.94).

**Figure S5:**
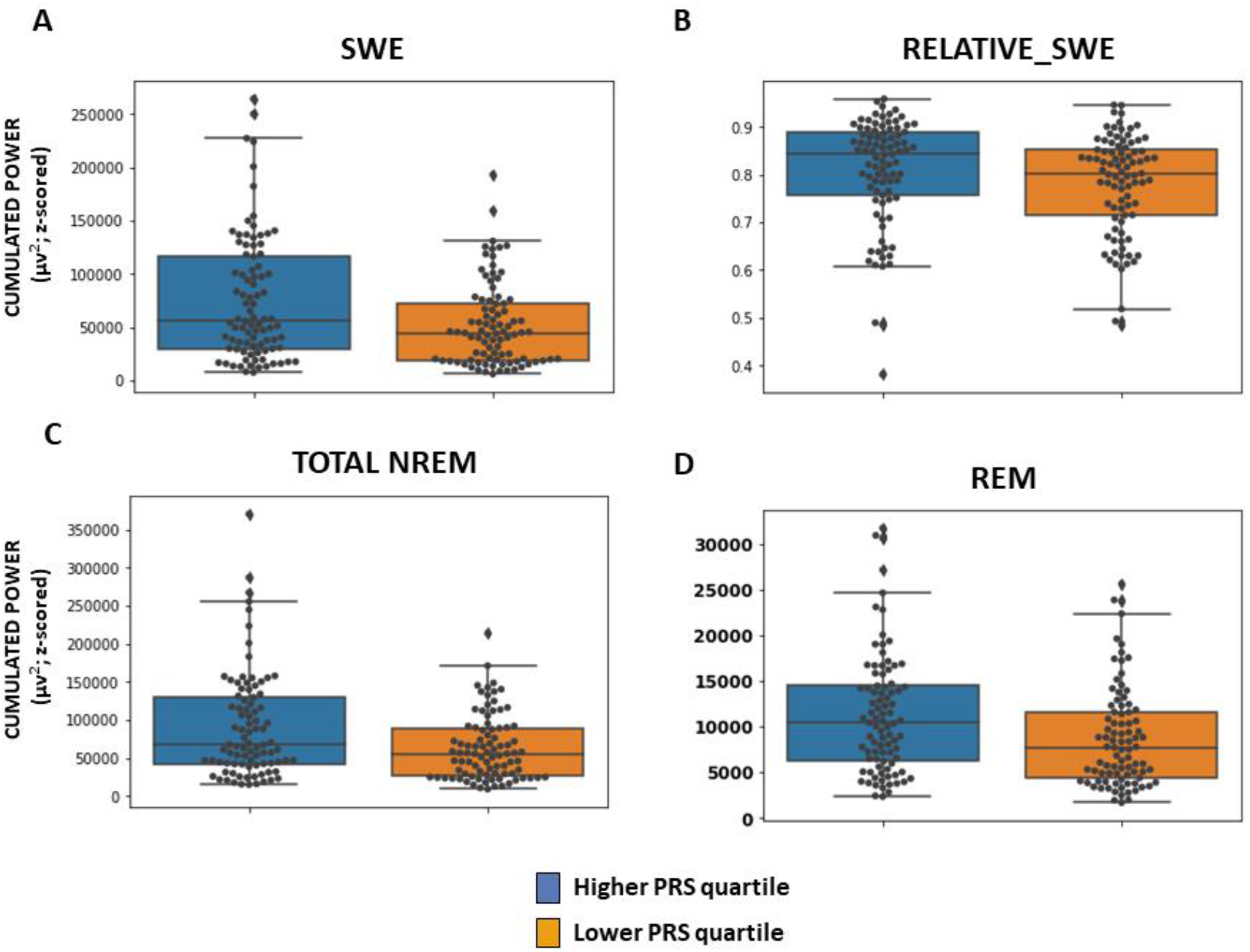
Differences in SWE, relative_SWE, total NREM power and REM power during baseline sleep between lower and upper AD PRS quartiles. Comparison of individual with 25% lowest (N = 90) and 25% highest (N = 90) PRS (using all SNPs) for SWE (**A**), relative SWE (**B**), cumulated total power between 0.5 and 25 Hz during NREM sleep (**C**) and cumulated power over the 2 to 6 Hz band during REM sleep (**D**). T-tests indicated higher power in higher quartile vs. lower quartile in all three absolute measures (**SWE: p = 0**.**008; total NREM: p = 0**.**008; REM: p = 0**.**01**) but not for **relative SWE (p = 0**.**14)**. Relative power was computed as the individual ratio between SWE and cumulated total power between 0.5 and 25 Hz during NREM sleep. Dots: individual values; boxes: interquartile interval; horizontal bar within boxes: median; error bars: 10 to 90% interval.

**Table S1.** Number of SNPs included in PRS computation as a function of p-value thresholding in reference GWAS summary statistic of ^21^.

| **P-value threshold** | **Number of SNPs included in PRS computation** |
| --- | --- |
| P-value threshold $5 \times 10^{-8}$ (and LD) | 64 |
| P-value threshold $10^{-6}$ (and LD) | 108 |
| P-value threshold $10^{-4}$ (and LD) | 517 |
| P-value threshold $10^{-3}$ (and LD) | 2543 |
| P-value threshold 0.01 (and LD) | 15654 |
| P-value threshold 0.05 (and LD) | 58096 |
| P-value threshold 0.1 (and LD) | 101336 |
| P-value threshold 0.3 (and LD) | 233635 |
| P-value threshold 0.5 (and LD) | 329341 |
| P-value threshold 1 (and LD) | 466829 |
| All SNPs (no LD) | 7780254 |

LD: linkage disequilibrium pruning

**Table S2.**
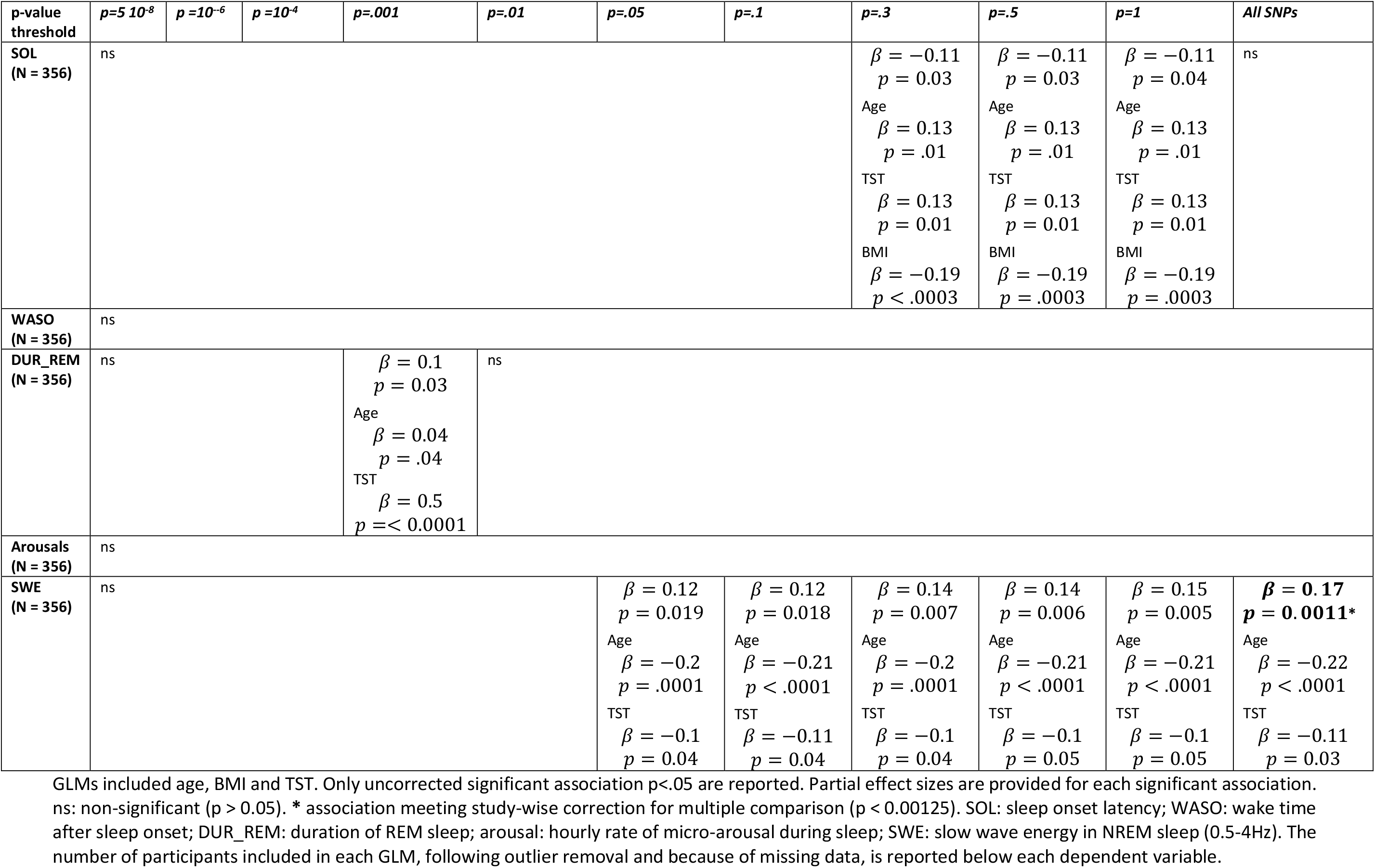
Statistical outcomes of GLMs with the five baseline night sleep metrics of interest vs. AD PRS from conservative (p < 5×10^−8^) to inclusive (p < 1) p-value threshold and selecting all SNPs.

**Table S3.** Statistical outcomes of GLMs with SWE during the first hour of sleep (SWE 1h) in baseline (BAS) nights of sleep and when separating slow oscillation SWE (SO-SWE) and fast oscillation SWE (FO-SWE) from conservative (p < 5×10^−8^) to inclusive (p < 1) p-value threshold and when selecting all SNPs.

| p-value threshold | $p=5 \times 10^{-8}$ | $p=10^{-6}$ | $p=10^{-4}$ | $p=.001$ | $p=.01$ | $p=.05$ | $p=.1$ | $p=.3$ | $p=.5$ | $p=1$ | All SNPs |
| --- | --- | --- | --- | --- | --- | --- | --- | --- | --- | --- | --- |
| **SWE 1h BAS (N = 355)** | ns | | | | | $\beta = 0.12$<br>$p = 0.02$<br>Age<br>$\beta = -0.2$<br>$p = .0001$<br>TST<br>$\beta = -0.17$<br>$p = 0.001$ | $\beta = 0.12$<br>$p = 0.03$<br>Age<br>$\beta = -0.21$<br>$p < .0001$<br>TST<br>$\beta = -0.17$<br>$p = 0.001$ | $\beta = 0.14$<br>$p = 0.005$<br>Age<br>$\beta = -0.20$<br>$p < .0001$<br>TST<br>$\beta = -0.17$<br>$p = 0.001$ | $\beta = 0.14$<br>$p = 0.005$<br>Age<br>$\beta = -0.21$<br>$p < .0001$<br>TST<br>$\beta = -0.17$<br>$p = 0.001$ | $\beta = 0.15$<br>$p = 0.004$<br>Age<br>$\beta = -0.21$<br>$p < .0001$<br>TST<br>$\beta = -0.17$<br>$p = 0.001$ | $\beta = 0.17$<br>$p = 0.0007$<br>Age<br>$\beta = -0.22$<br>$p < .0001$<br>TST<br>$\beta = -0.17$<br>$p = 0.0007$ |
| **SO-SWE (N = 357)** | ns | | | | | $\beta = 0.11$<br>$p = 0.04$<br>Age<br>$\beta = -0.2$<br>$p = .0002$<br>TST<br>$\beta = -0.14$<br>$p = 0.007$ | $\beta = 0.12$<br>$p = 0.02$<br>Age<br>$\beta = -0.2$<br>$p = .0001$<br>TST<br>$\beta = -0.14$<br>$p = 0.007$ | $\beta = 0.12$<br>$p = 0.02$<br>Age<br>$\beta = -0.2$<br>$p = .0001$<br>TST<br>$\beta = -0.14$<br>$p = 0.007$ | $\beta = 0.12$<br>$p = 0.02$<br>Age<br>$\beta = -0.2$<br>$p = .0001$<br>TST<br>$\beta = -0.14$<br>$p = 0.007$ | $\beta = 0.12$<br>$p = 0.2$<br>Age<br>$\beta = -0.2$<br>$p = .0001$<br>TST<br>$\beta = -0.14$<br>$p = 0.007$ | $\beta = 0.16$<br>$p = 0.003$<br>Age<br>$\beta = -0.22$<br>$p < .0001$<br>TST<br>$\beta = -0.14$<br>$p = 0.007$ |
| **FO-SWE (N = 357)** | ns | | | | | $\beta = 0.11$<br>$p = 0.03$<br>Age<br>$\beta = -0.2$<br>$p = .0002$<br>TST<br>$\beta = -0.14$<br>$p = 0.007$ | $\beta = 0.11$<br>$p = 0.04$<br>Age<br>$\beta = -0.2$<br>$p = .0001$<br>TST<br>$\beta = -0.14$<br>$p = 0.007$ | $\beta = 0.12$<br>$p = 0.02$<br>Age<br>$\beta = -0.2$<br>$p = .0001$<br>TST<br>$\beta = -0.14$<br>$p = 0.007$ | $\beta = 0.12$<br>$p = 0.02$<br>Age<br>$\beta = -0.2$<br>$p = .0001$<br>TST<br>$\beta = -0.14$<br>$p = 0.007$ | $\beta = 0.13$<br>$p = 0.01$<br>Age<br>$\beta = -0.2$<br>$p = .0001$<br>TST<br>$\beta = -0.14$<br>$p = 0.007$ | $\beta = 0.15$<br>$p = 0.005$<br>Age<br>$\beta = -0.21$<br>$p < .0001$<br>TST<br>$\beta = -0.14$<br>$p = 0.007$ |

GLMs included age and BMI (and TST for SO-SWE and FO-SWE). Only uncorrected significant association $p < .05$ are reported. Partial effect sizes are provided for each significant association. ns: non-significant ( $p > 0.05$ ). BMI was never significantly associated with AD PRS. SO-SWE: 0.5-1Hz; FO-SWE: 1.25-4Hz.

**Table S4.** Statistical outcomes of GLMs with SWE during recovery and extension and with SWE rebound vs. AD PRS from conservative (p < 5×10^−8^) to inclusive (p < 1) p-value threshold and selecting all SNPs.

| *P-value threshold* | *p=5 10<sup>-8</sup>* | *p=10<sup>-6</sup>* | *p=10<sup>-4</sup>* | *p=.001* | *p=.01* | *p=.05* | *p=.1* | *p=.3* | *p=.5* | *p=1* | *All SNPs* |
| --- | --- | --- | --- | --- | --- | --- | --- | --- | --- | --- | --- |
| **SWE – REC**<br>**(N = 353)** | ns | | | | | | $\beta = 0.11$<br>$p = 0.04$ | $\beta = 0.13$<br>$p = 0.008$ | $\beta = 0.13$<br>$p = 0.008$ | $\beta = 0.14$<br>**$p = 0.006^*$** | $\beta = 0.11$<br>$p = 0.04$ |
| | | | | | | | Age<br>$\beta = -0.28$<br>$p < .0001$ | Age<br>$\beta = -0.28$<br>$p < .0001$ | Age<br>$\beta = -0.29$<br>$p < .0001$ | Age<br>$\beta = -0.29$<br>$p < .0001$ | Age<br>$\beta = -0.3$<br>$p < .0001$ |
| **SWE 1h REC**<br>**(N = 355)** | ns | | | | | | $\beta = 0.11$<br>$p = 0.03$ | $\beta = 0.16$<br>$p = 0.002$ | $\beta = 0.16$<br>$p = 0.002$ | $\beta = 0.16$<br>$p = 0.002$ | $\beta = 0.12$<br>$p = 0.02$ |
| | | | | | | | Age<br>$\beta = -0.27$<br>$p < .0001$ | Age<br>$\beta = -0.27$<br>$p < .0001$ | Age<br>$\beta = -0.27$<br>$p < .0001$ | Age<br>$\beta = -0.27$<br>$p < .0001$ | Age<br>$\beta = -0.28$<br>$p < .0001$ |
| **SWE – Rebound**<br>**(N = 344)** | ns | | | | | | | | | | $\beta = -0.11$<br>$p = 0.047$ |
| **SWE – EXT**<br>**(N = 356)** | ns |  |  |  |  |  |  |  |  |  |  |

GLMs included age, BMI and total sleep time (TST). Only uncorrected significant association $p < 0.05$ are reported (in bold). \* association meeting study-wise correction for multiple comparison ( $p < 0.00625$ ). SWE rebound consist in the difference between difference in the SWE in the first hour of sleep of recovery and baseline nights. Partial effect sizes are provide for each significant association. ns: non-significant ( $p > 0.05$ ). The number of participants included in each GLM, following outlier removal and because of missing data, is reported below each dependent variable.

**Table S5.** Statistical outcomes of GLMs with non-EEG sleep metrics vs. AD PRS from conservative (p < 5×10^−8^) to inclusive (p < 1) p-value threshold and selecting all SNPs.

| *p-value threshold* | *p=5 10<sup>-8</sup>* | *p=10<sup>-6</sup>* | *p=10<sup>-4</sup>* | *p=.001* | *p=.01* | *p=.05* | *p=.1* | *p=.3* | *p=.5* | *p=1* | *All SNPs* |
| --- | --- | --- | --- | --- | --- | --- | --- | --- | --- | --- | --- |
| **kRA**<br>(N = 361) | $\beta = -0.11$<br>$p = 0.036$ | $\beta = -0.13$<br>$p = 0.016$ | ns | ns | ns | ns | ns | ns | ns | ns | ns |
| **Sleep-qual**<br>(N = 363) | ns | ns | ns | ns | ns | ns | ns | ns | ns | ns | ns |
| **Day-sleepiness**<br>(N = 363) | ns | ns | $\beta = 0.11$<br>$p = 0.043$ | $\beta = 0.12$<br>$p = 0.02$ | $\beta = 0.13$<br>$p = 0.01$ | **$\beta = 0.17$**<br>**$p = 0.001$ \*** | $\beta = 0.16$<br>$p = 0.003$ | **$\beta = 0.16$**<br>**$p = 0.002$ \*** | $\beta = 0.16$<br>$p = 0.003$ | $\beta = 0.15$<br>$p = 0.006$ | ns |

GLMs included age and BMI. Only uncorrected significant association $p < .05$ are reported (in bold). Partial effect sizes are provided for each significant association. ns: non-significant ( $p > 0.05$ ) BMI and age were not significantly associated with the dependent variables. \* associations meeting study-wise correction for multiple comparison ( $p < 0.002$ ). The number of participants included in each GLM, following outlier removal and because of missing data, is reported below each dependent variable

## References

1. Musiek ES, Holtzman DM. Three dimensions of the amyloid hypothesis: Time, space and “wingmen.” Nat Neurosci. 2015;18(6):800–806. doi:10.1038/nn.4018

2. Jack CR, Bennett DA, Blennow K, et al. NIA-AA Research Framework: Toward a biological definition of Alzheimer’s disease. Alzheimer’s Dement. 2018;14(4):535–562. doi:10.1016/j.jalz.2018.02.018

3. Scheltens P, Blennow K, Breteler MMB, et al. Alzheimer’s disease. Lancet. 2016;388(10043):505–517. doi:10.1016/S0140-6736(15)01124-1

4. Norton S, Matthews FE, Barnes DE, Yaffe K, Brayne C. Potential for primary prevention of Alzheimer’s disease: An analysis of population-based data. Lancet Neurol. 2014;13(8):788–794. doi:10.1016/S1474-4422(14)70136-X

5. Van Egroo M, Narbutas J, Chylinski D, et al. Sleep–wake regulation and the hallmarks of the pathogenesis of Alzheimer’s disease. Sleep. 2019;42(4). doi:10.1093/sleep/zsz017

6. Al-Qassabi A, Fereshtehnejad SM, Postuma RB. Sleep Disturbances in the Prodromal Stage of Parkinson Disease. Curr Treat Options Neurol. 2017;19(6):22. doi:10.1007/s11940-017-0458-1

7. Mander BA, Marks SM, Vogel JW, et al. β-amyloid disrupts human NREM slow waves and related hippocampus-dependent memory consolidation. Nat Neurosci. 2015;18(7):1051–1057. doi:10.1038/nn.4035

8. Lucey BP, McCullough A, Landsness EC, et al. Reduced non–rapid eye movement sleep is associated with tau pathology in early Alzheimer’s disease. Sci Transl Med. 2019;11(474):eaau6550. doi:10.1126/SCITRANSLMED.AAU6550

9. Branger P, Arenaza-Urquijo EM, Tomadesso C, et al. Relationships between sleep quality and brain volume, metabolism, and amyloid deposition in late adulthood. Neurobiol Aging. 2016;41:107–114. doi:10.1016/j.neurobiolaging.2016.02.009

10. Pase MP, Himali JJ, Grima NA, et al. Sleep architecture and the risk of incident dementia in the community. Neurology. 2017;89(12):10.1212/WNL.0000000000004373. doi:10.1212/WNL.0000000000004373

11. Lim ASP, Kowgier M, Yu L, Buchman AS, Bennett DA. Sleep Fragmentation and the Risk of Incident Alzheimer’s Disease and Cognitive Decline in Older Persons. Sleep. 2013;36(7):1027–1032. doi:10.5665/sleep.2802

12. Holth JK, Fritschi SK, Wang C, et al. The sleep-wake cycle regulates brain interstitial fluid tau in mice and CSF tau in humans. Science (80-). January 2019:eaav2546. doi:10.1126/science.aav2546

13. Ooms S, Overeem S, Besse K, Rikkert MO, Verbeek M, Claassen J a HR. Effect of 1 Night of Total Sleep Deprivation on Cerebrospinal Fluid β-Amyloid 42 in Healthy Middle-Aged Men: A Randomized Clinical Trial. JAMA Neurol. 2014;71(8):971–977. doi:10.1001/jamaneurol.2014.1173

14. Ju Y-ES, Ooms SJ, Sutphen C, et al. Slow wave sleep disruption increases cerebrospinal fluid amyloid-β levels. Brain. 2017;140(8):2104–2111. doi:10.1093/brain/awx148

15. Mather M, Harley CW. The Locus Coeruleus: Essential for Maintaining Cognitive Function and the Aging Brain. Trends Cogn Sci. 2016;20(3):214–226. doi:10.1016/j.tics.2016.01.001

16. Braak H, Del Tredici K. The pathological process underlying Alzheimer’s disease in individuals under thirty. Acta Neuropathol. 2011;121(2):171–181. doi:10.1007/s00401-010-0789-4

17. Gatz M, Reynolds CA, Fratiglioni L, et al. Role of genes and environments for explaining Alzheimer disease. Arch Gen Psychiatry. 2006;63(2):168–174. doi:10.1001/archpsyc.63.2.168

18. Ertekin-Taner N. Genetics of Alzheimer disease in the pre- and post-GWAS era. Alzheimer’s Res Ther. 2010;2(1):3. doi:10.1186/alzrt26

19. Euesden J, Lewis CM, O’Reilly PF. PRSice: Polygenic Risk Score software. Bioinformatics. 2015;31(9):1466–1468. doi:10.1093/bioinformatics/btu848

20. Ge T, Chen C-Y, Ni Y, Feng Y-CA, Smoller JW. Polygenic prediction via Bayesian regression and continuous shrinkage priors. Nat Commun. 2019;10(1):1776. doi:10.1038/s41467-019-09718-5

21. Martiskainen H, Helisalmi S, Viswanathan J, et al. Effects of Alzheimer’s disease-associated risk loci on cerebrospinal fluid biomarkers and disease progression: A polygenic risk score approach. J Alzheimer’s Dis. 2015;43(2):565–573. doi:10.3233/JAD-140777

22. Ge T, Sabuncu MR, Smoller JW, Sperling RA, Mormino EC. Dissociable influences of APOE ε4 and polygenic risk of AD dementia on amyloid and cognition. Neurology. 2018;90(18):e1605–e1612. doi:10.1212/WNL.0000000000005415

23. Sabuncu MR, Buckner RL, Smoller JW, Lee PH, Fischl B, Sperling RA. The association between a polygenic Alzheimer score and cortical thickness in clinically normal subjects. Cereb Cortex. 2012;22(11):2653–2661. doi:10.1093/cercor/bhr348

24. Marden JR, Mayeda ER, Walter S, et al. Using an Alzheimer disease polygenic risk score to predict memory decline in black and white Americans over 14 years of follow-up. Alzheimer Dis Assoc Disord. 2016;30(3):195–202. doi:10.1097/WAD.0000000000000137

25. Mormino EC, Sperling RA, Holmes AJ, et al. Polygenic risk of Alzheimer disease is associated with early- and late-life processes. Neurology. 2016;87(5):481–488. doi:10.1212/WNL.0000000000002922

26. Foley SF, Tansey KE, Caseras X, et al. Multimodal Brain Imaging Reveals Structural Differences in Alzheimer’s Disease Polygenic Risk Carriers: A Study in Healthy Young Adults. Biol Psychiatry. 2017;81(2):154–161. doi:10.1016/j.biopsych.2016.02.033

27. Beck AT, Steer RA, G. Gm. Psychometric Properties of the Beck Depression Inventory: Twenty-five years of evaluation. Clin Psychol Rev. 1988;8:77–100. doi:https://doi.org/10.1016/0272-7358(88)90050-5

28. Buysse DJ, Reynolds CF, Monk TH, Berman SR, Kupfer DJ. The Pittsburgh sleep quality index: A new instrument for psychiatric practice and research. Psychiatry Res. 1989;28(2):193–213. doi:10.1016/0165-1781(89)90047-4

29. Johns MW. A new method for measuring daytime sleepiness: The Epworth Sleepiness Scale. Sleep. 1991;14(6):540–545. doi:10.1016/j.sleep.2007.08.004

30. John, Raven J. Raven Progressive Matrices. In: Handbook of Nonverbal Assessment. Boston, MA: Springer US; 2003:223–237. doi:10.1007/978-1-4615-0153-4_11

31. Berthomier C, Muto V, Schmidt C, et al. Exploring scoring methods for research studies?: Accuracy and variability of visual and automated sleep scoring. 2020;(August 2019):1–11. doi:10.1111/jsr.12994

32. Wallant DC t., Muto V, Gaggioni G, et al. Automatic artifacts and arousals detection in whole-night sleep EEG recordings. J Neurosci Methods. 2016;258:124–133. doi:10.1016/j.jneumeth.2015.11.005

33. Skorucak J, Arbon EL, Dijk D-J, Achermann P. Response to chronic sleep restriction, extension, and total sleep deprivation in humans: adaptation or preserved sleep homeostasis? Sleep. 2018;(May):1–27. doi:10.1093/ntr/ntx198/4104547/Differences-in-adolescent-e-cigarette-and

34. Schmidt C, Peigneux P, Cajochen C. Age-related changes in sleep and circadian rhythms: Impact on cognitive performance and underlying neuroanatomical networks. Front Neurol. 2012;JUL:118. doi:10.3389/fneur.2012.00118

35. Dijk D-J, Landolt H-P. Sleep Physiology, Circadian Rhythms, Waking Performance and the Development of Sleep-Wake Therapeutics. In: Handbook of Experimental Pharmacology. Springer, Berlin, Heidelberg; 2019:1–41. doi:10.1007/164_2019_243

36. Purcell S, Neale B, Todd-Brown K, et al. PLINK: a tool set for whole-genome association and population-based linkage analyses. Am J Hum Genet. 2007;81(3):559–575. doi:10.1086/519795

37. Altshuler DL, Durbin RM, Abecasis GR, et al. A map of human genome variation from population-scale sequencing. Nature. 2010;467(7319):1061–1073. doi:10.1038/nature09534

38. Yengo L, Sidorenko J, Kemper KE, et al. Meta-analysis of genome-wide association studies for height and body mass index in ∼700000 individuals of European ancestry. Hum Mol Genet. August 2018. doi:10.1093/hmg/ddy271

39. Marioni RE, Harris SE, Zhang Q, et al. GWAS on family history of Alzheimer’s disease. Transl Psychiatry. 2018;8(1):99. doi:10.1038/s41398-018-0150-6

40. Sudlow C, Gallacher J, Allen N, et al. UK biobank: an open access resource for identifying the causes of a wide range of complex diseases of middle and old age. PLoS Med. 2015;12(3):e1001779. doi:10.1371/journal.pmed.1001779

41. Lambert JC, Ibrahim-Verbaas CA, Harold D, et al. Meta-analysis of 74,046 individuals identifies 11 new susceptibility loci for Alzheimer’s disease. Nat Genet. 2013;45(12):1452–1458. doi:10.1038/ng.2802

42. Sleegers K, Bettens K, De Roeck A, et al. A 22-single nucleotide polymorphism Alzheimer’s disease risk score correlates with family history, onset age, and cerebrospinal fluid Aβ42. Alzheimers Dement. 2015;11(12):1452–1460. doi:10.1016/j.jalz.2015.02.013

43. Escott-Price V, Sims R, Bannister C, et al. Common polygenic variation enhances risk prediction for Alzheimer’s disease. Brain. 2015;138(12):3673–3684. doi:10.1093/brain/awv268

44. Hammad G, Reyt M. ghammad/pyActigraphy: Actigraphy made simple! January 2019. doi:10.5281/ZENODO.2537921

45. Faul F, Erdfelder E, Lang A-G, Buchner A.G * Power 3: A flexible statistical power analysis program for the social, behavioral, and biomedical sciences. 2007;39(2):175–191.

46. Santoro ML, Ota V, de Jong S, et al. Polygenic risk score analyses of symptoms and treatment response in an antipsychotic-naive first episode of psychosis cohort. Transl Psychiatry. 2018;8(1):174. doi:10.1038/s41398-018-0230-7

47. Dudbridge F. Power and Predictive Accuracy of Polygenic Risk Scores. Wray NR, ed. PLoS Genet. 2013;9(3):e1003348. doi:10.1371/journal.pgen.1003348

48. Ettore E, Bakardjian H, Solé M, et al. Relationships between objectives sleep parameters and brain amyloid load in subjects at risk to Alzheimer’s disease: the INSIGHT-preAD Study. Sleep. July 2019. doi:10.1093/sleep/zsz137

49. Carrier J, Viens I, Poirier G, et al. Sleep slow wave changes during the middle years of life. Eur J Neurosci. 2011;33(4):758–766. doi:10.1111/j.1460-9568.2010.07543.x

50. Kunkle BW, Grenier-Boley B, Sims R, et al. Genetic meta-analysis of diagnosed Alzheimer’s disease identifies new risk loci and implicates Aβ, tau, immunity and lipid processing. Nat Genet. 2019;51(3):414–430. doi:10.1038/s41588-019-0358-2

51. Dijk DJ, Czeisler C a. Contribution of the circadian pacemaker and the sleep homeostat to sleep propensity, sleep structure, electroencephalographic slow waves, and sleep spindle activity in humans. J Neurosci. 1995;15(5 Pt 1):3526–3538. http://www.ncbi.nlm.nih.gov/entrez/query.fcgi?cmd=Retrieve&db=PubMed&dopt=Citation&list_uids=7751928.

52. Steriade M, Amzica F. Slow sleep oscillation, rhythmic K-complexes, and their paroxysmal developments. J Sleep Res. 1998;7(S1):30–35. doi:10.1046/j.1365-2869.7.s1.4.x

53. Klerman EB, Dijk DJ. Interindividual variation in sleep duration and its association with sleep debt in young adults. Sleep. 2005;28(10):1253–1259. doi:10.1093/sleep/28.10.1253

54. Viola AU, Archer SN, James LMM, et al. PER3 Polymorphism Predicts Sleep Structure and Waking Performance. Curr Biol. 2007;17(7):613–618. doi:10.1016/j.cub.2007.01.073

55. Tononi G, Cirelli C. Sleep and the Price of Plasticity: From Synaptic and Cellular Homeostasis to Memory Consolidation and Integration. Neuron. 2014;81(1):12–34. doi:10.1016/j.neuron.2013.12.025

56. Scammell TE, Arrigoni E, Lipton JO. Neural Circuitry of Wakefulness and Sleep. Neuron. 2017;93(4):747–765. doi:10.1016/j.neuron.2017.01.014

57. Huber R, Mäki H, Rosanova M, et al. Human cortical excitability increases with time awake. Cereb Cortex. 2013;23(2):332–338. doi:10.1093/cercor/bhs014

58. Ly JQM, Gaggioni G, Chellappa SL, et al. Circadian regulation of human cortical excitability. Nat Commun. 2016;In Press:11828. doi:10.1038/ncomms11828

59. Dash MB, Douglas CL, Vyazovskiy V V., Cirelli C, Tononi G. Long-Term Homeostasis of Extracellular Glutamate in the Rat Cerebral Cortex across Sleep and Waking States. J Neurosci. 2009;29(3):620–629. doi:10.1523/JNEUROSCI.5486-08.2009

60. Hefti K, Holst SC, Sovago J, et al. Increased metabotropic glutamate receptor subtype 5 availability in human brain after one night without sleep. Biol Psychiatry. 2013;73(2):161–168. doi:10.1016/j.biopsych.2012.07.030

61. Bero AW, Yan P, Roh JH, et al. Neuronal activity regulates the regional vulnerability to amyloid-ß deposition. Nat Neurosci. 2011;14(6):750–756. doi:10.1038/nn.2801

62. Bero AW, Bauer AQ, Stewart FR, et al. Bidirectional relationship between functional connectivity and amyloid-β deposition in mouse brain. J Neurosci. 2012;32(13):4334–4340. doi:10.1523/JNEUROSCI.5845-11.2012

63. Thal D, Rüb U, Orantes M, Braak H. Phases of A beta-deposition in the human brain and its relevance for the development af AD. Neurology. 2002;58:1791–1800.

64. Braak H, Del Tredici K. The preclinical phase of the pathological process underlying sporadic Alzheimer’s disease. Brain. 2015;138(10):2814–2833. doi:10.1093/brain/awv236

65. Roh JH, Huang Y, Bero a. W, et al. Disruption of the Sleep-Wake Cycle and Diurnal Fluctuation of β-Amyloid in Mice with Alzheimer’s Disease Pathology. Sci Transl Med. 2012;4(150):150ra122–150ra122. doi:10.1126/scitranslmed.3004291

66. Chai X, Dage JL, Citron M. Constitutive secretion of tau protein by an unconventional mechanism. Neurobiol Dis. 2012;48(3):356–366. doi:10.1016/j.nbd.2012.05.021

67. Yamada K, Holth JK, Liao F, et al. Neuronal activity regulates extracellular tau in vivo. J Exp Med. 2014;211(3):387–393. doi:10.1084/jem.20131685

68. Schultz MK, Gentzel R, Usenovic M, et al. Pharmacogenetic neuronal stimulation increases human tau pathology and trans-synaptic spread of tau to distal brain regions in mice. Neurobiol Dis. 2018;118(June):161–176. doi:10.1016/j.nbd.2018.07.003

69. Crimins JL, Rocher AB, Luebke JI. Electrophysiological changes precede morphological changes to frontal cortical pyramidal neurons in the rTg4510 mouse model of progressive tauopathy. Acta Neuropathol. 2012;124(6):777–795. doi:10.1007/s00401-012-1038-9

70. Nilsen LH, Rae C, Ittner LM, Götz J, Sonnewald U. Glutamate metabolism is impaired in transgenic mice with tau hyperphosphorylation. J Cereb Blood Flow Metab. 2013;33(5):684–691. doi:10.1038/jcbfm.2012.212

71. Holth JK, Bomben VC, Reed JG, et al. Tau Loss Attenuates Neuronal Network Hyperexcitability in Mouse and Drosophila Genetic Models of Epilepsy. J Neurosci. 2013;33(4):1651–1659. doi:10.1523/jneurosci.3191-12.2013

72. Polydoro M, Dzhala VI, Pooler AM, et al. Soluble pathological tau in the entorhinal cortex leads to presynaptic deficits in an early Alzheimer’s disease model. Acta Neuropathol. 2014;127(2):257–270. doi:10.1007/s00401-013-1215-5

73. Van der Jeugd A, Hochgräfe K, Ahmed T, et al. Cognitive defects are reversible in inducible mice expressing pro-aggregant full-length human Tau. Acta Neuropathol. 2012;123(6):787–805. doi:10.1007/s00401-012-0987-3

74. Sydow A, Van der Jeugd A, Zheng F, et al. Tau-induced defects in synaptic plasticity, learning, and memory are reversible in transgenic mice after switching off the toxic Tau mutant. J Neurosci. 2011;31(7):2511–2525. doi:10.1523/JNEUROSCI.5245-10.2011

75. Oddo S, Caccamo A, Shepherd JD, et al. Triple-Transgenic Model of Alzheimer’s Disease with Plaques and Tangles. Neuron. 2003;39(3):409–421. doi:10.1016/S0896-6273(03)00434-3

76. Fein JA, Sokolow S, Miller CA, et al. Co-localization of amyloid beta and tau pathology in Alzheimer’s disease synaptosomes. Am J Pathol. 2008;172(6):1683–1692. doi:10.2353/ajpath.2008.070829

77. Svetnik V, Snyder ES, Ma J, Tao P, Lines C, Herring WJ. EEG spectral analysis of NREM sleep in a large sample of patients with insomnia and good sleepers: effects of age, sex and part of the night. J Sleep Res. 2017;26(1):92–104. doi:10.1111/jsr.12448

78. Horne JA, Ostberg O. A self-assessment questionnaire to determine morningness-eveningness in human circadian rhythms. Int J Chronobiol. 1976;4(2):97–110. http://www.ncbi.nlm.nih.gov/pubmed/1027738. accessed May 18, 2017.

